# A systematic review of resting state functional MRI connectivity changes and cognitive impairment in multiple sclerosis

**DOI:** 10.1101/2021.03.18.21253878

**Authors:** Danka Jandric, Anisha Doshi, Richelle Scott, David Paling, David Rog, Jeremy Chataway, Menno Schoonheim, Geoff Parker, Nils Muhlert

**Affiliations:** Division of Neuroscience & Experimental Psychology, School of Biological Sciences, Faculty of Biology, Medicine and Health, University of Manchester, Manchester Academic Health Science Centre, Manchester, UK; Queen Square Multiple Sclerosis Centre, Department of Neuroinflammation, UCL Institute of Neurology, Faculty of Brain Sciences, University College London, UK; Royal Hallamshire Hospital, Sheffield Teaching Hospitals, NHS UK; Salford Royal Hospital, Salford Royal NHS Foundation Trust, NHS UK; Department of Anatomy and Neurosciences, MS Center Amsterdam, Amsterdam Neuroscience, Amsterdam UMC, Vrije Universiteit Amsterdam, Amsterdam, The Netherlands.; Centre for Medical Image Computing, Department of Computer Science and Department of Neuroinflammation, Queen Square Institute of Neurology, University College London, London, UK

## Abstract

Cognitive impairment is common in multiple sclerosis (MS) and resting state functional MRI (rs-fMRI) functional connectivity (FC) is increasingly used to study its pathophysiology. However, results remain difficult to interpret, showing both high and low FC associated with cognitive impairment. We conducted a systematic review of rs-fMRI studies in MS to understand whether the direction of FC change relates to cognitive dysfunction, and how this may be influenced by the choice of methodology. Embase, Medline and PsycINFO were searched for studies assessing cognitive function and rs-fMRI FC in adults with MS. Fifty-seven studies were included in a narrative synthesis. Of these, 50 found an association between cognitive impairment and FC abnormalities. Worse cognition was linked to high FC in 18 studies, and to low FC in 17 studies. Nine studies found patterns of both high and low FC related to poor cognitive performance, in different regions or for different MR metrics. There was no clear link to increased FC during early stages of MS and reduced FC in later stages, as predicted by common models of MS pathology. Throughout, we found substantial heterogeneity in study methodology, and carefully consider how this may impact on the observed findings. These results indicate an urgent need for greater standardisation in the field – in the choice of MRI analysis and the definition of cognitive impairment. Through this we will be closer to using rs-fMRI FC as a biomarker in clinical studies, and as a tool to understand mechanisms underpinning cognitive symptoms in MS.

**Key points:**

- Cognitive impairment in multiple sclerosis (MS) is increasingly being researched with advanced magnetic resonance imaging (MRI) measures, including resting state functional MRI (rs-fMRI)
- The rs-fMRI functional connectivity (FC) metric is associated with cognitive impairment, and has the potential to be a biomarker of cognitive decline.
- A main challenge to developing a FC biomarker is the lack of consistency in the direction of FC changes associated with cognitive impairment, with cognitive deficits associated with both lower and higher FC.
- FC changes don’t appear to be linked to clinical and methodological factors such as disease phenotype, disease duration and brain region or network studied.
- Overall, there is substantial heterogeneity in study methodology, suggesting an acute need to standardise the study of cognitive impairment in MS and its investigation by rs-fMRI methods.

## Introduction

Multiple sclerosis (MS) is a chronic immune mediated disorder of the central nervous system that predominantly affects young adults (1–3). Inflammatory demyelination is pathognomonic with neurodegeneration insidiously dominating over time (4).

Cognitive impairment is common in all MS phenotypes (5–7) with an estimated prevalence of 43-70% dependent on factors including phenotype and the cognitive diagnostic criteria used (8, 9). Cognitive impairment is associated with several adverse outcomes including a higher risk of depression, unemployment and reduced quality of life (9–11). A more progressive MS phenotype and longer disease duration have been shown to be associated with greater cognitive impairment (12–16). There are currently no licensed treatments for cognitive symptoms in MS, however exercise (17) and behavioural therapy show promise (18). Disease modifying therapies show positive outcomes on cognitive dysfunction in MS, despite no routine evaluation in phase 3 clinical trials currently. However, effects are small and at present understudied, and there are to date no approved pharmaceutical treatments for cognitive symptoms (7, 19).

Gaining an understanding of the underlying pathophysiology of cognitive dysfunction is essential for diagnosing, monitoring and developing treatments for this debilitating aspect of MS. The ‘clinico-radiological’ paradox highlights the mismatch of MS cognitive symptoms and conventional Magnetic Resonance Imaging (MRI) measures, such as lesion volumes (20). It is widely accepted that cognitive function is supported by a complex network of structurally interconnected brain regions supporting a highly dynamic functional network, which is researched with advanced MRI tools such as resting state functional MRI (rs-fMRI), in MS and other neurodegenerative diseases (20–24).

The main measure derived from rs-fMRI is the functional connectivity (FC) metric. It is a measure of the statistical correlation of blood-oxygenation-level-dependent (BOLD) signal time course between any selection of voxels. The underlying assumption is that voxels with similar BOLD time courses are connected in the performance of a function (25), see Figure 1. FC has the potential to be an imaging biomarker of cognitive performance in neurodegenerative disease (26) and is the subject of a growing research field in MS (7). Such a marker could offer a fast, non-invasive way to detect imminent cognitive decline, which is often underdiagnosed on routine neurological examinations (27). For a measure to be suitable as a clinical biomarker, it needs to be able to identify those with cognitive dysfunction from those without it, and to show acceptable repeatability and reproducibility across studies. In some diseases, like Alzheimer’s disease, the rs-fMRI literature shows consistently low FC in the default mode network (DMN) (28), yet a recent review of rs-fMRI studies in several neurodegenerative diseases, including Alzheimer’s, argued that the evidence is not yet strong enough for rs-fMRI FC measures to be suitable biomarkers (26). This review cited a lack of standardised protocols as a challenge in the field.

**Figure 1.**
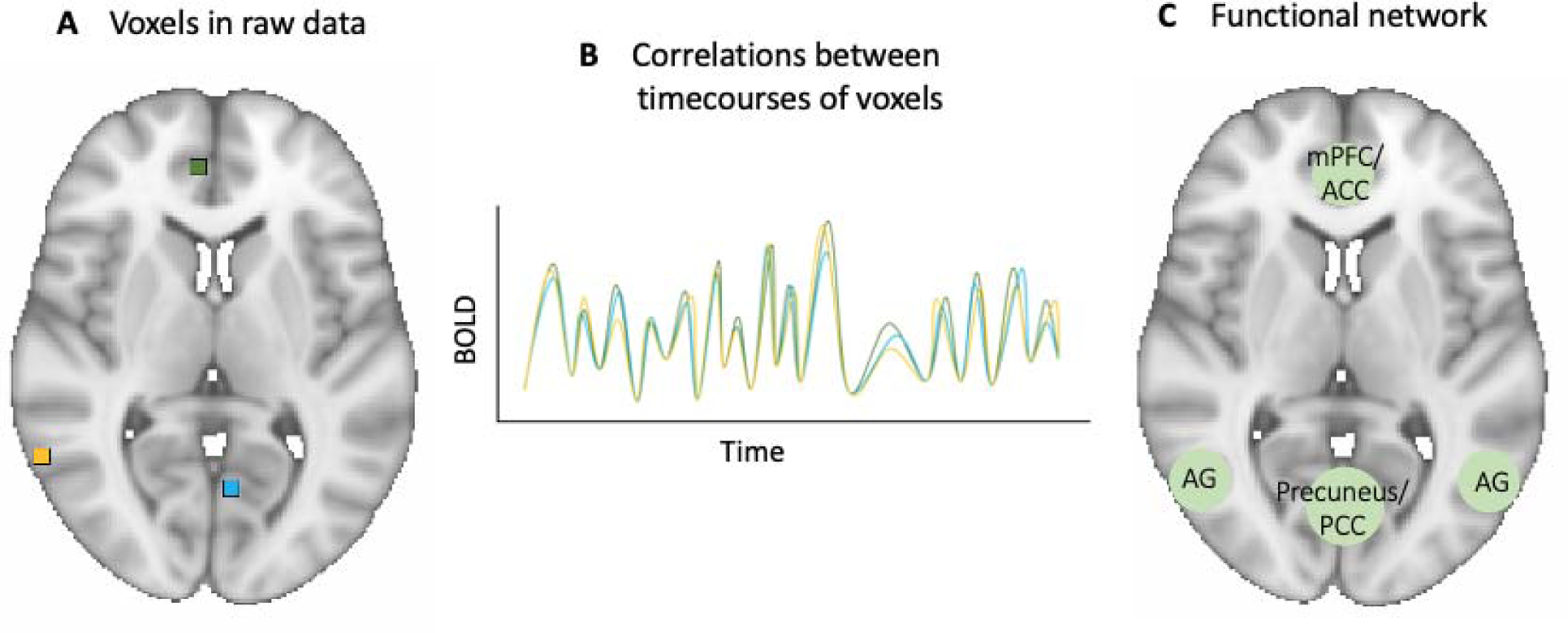
Schematic of functional connectivity and a functional network. Functional connectivity is a measure of the statistical correlation of blood-oxygenation-level-dependent signal timecourses (part **B**) between any selection of voxels (part **A**). Voxels or voxel clusters showing high correlations are considered functionally connected, and can be used to identify functional networks such as the default mode network (part **C**). Abbreviations: ACC = anterior cingulate cortex; AG = angular gyrus; BOLD = Blood-oxygenation-level-dependent signal; mPFC = medial prefrontal cortex; PCC = posterior cingulate cortex

The rs-fMRI FC literature on cognition in MS has not yet been subject to systematic review, and so the specificity and reliability of FC as a marker of cognitive dysfunction has not been established. Correlations between FC metrics and cognition have been frequently reported (29–32), but in studies comparing FC between cognitively impaired (CI) and cognitively preserved (CP) patients, results have shown both high and low FC linked with worse cognitive function (33–37). A common interpretation of increases in any type of brain function is that of functional “reorganisation”: a compensatory mechanism that enables the functioning of networks in the presence of structural damage, hence delaying clinical progression. This compensatory mechanism is thought to be sustainable only up to a critical point, at which the structural damage becomes too great to compensate for, leading to the hypothesized “network collapse”, manifested as decreases in FC and clinical progression (38, 39). In support of this, several studies indicate different patterns of FC changes at different disease stages, such as high FC in clinically isolated syndrome (CIS), the earliest stage of MS, and low FC in progressive MS (35,40–43). However, high FC has also been related to the severity of impairment (29), casting doubt on the beneficial nature of these changes. As such, it is not yet clear whether the pattern of results from rs-fMRI studies consistently fits the predictions of this model. This may be complicated by the heterogeneity in methodological aspects of studies which could influence the direction of findings (44).

In this study we carry out a systematic review of rs-fMRI FC studies of cognitive dysfunction in MS to outline the state of the field and provide a critical analysis of findings to date. We considered directionality of results and the influence of methodological aspects on findings of FC alterations. Through doing so we offer key points that need to be addressed in order to develop a parsimonious account of why FC may change in MS and what it may mean for clinical practice.

## Method

### Protocol and Registration

The design of the systematic review and manuscript preparation were based on the Preferred Reporting Items for Systematic Reviews and Meta-Analysis (PRISMA) guidelines (45). The systematic review protocol was developed in advance and, in accordance with PRISMA guidelines, registered with the International Prospective Register of Systematic Reviews (PROSPERO) on 18 May 2020, and last updated on 31/8/2020 (registration number CRD42020154415).

### Information sources and search strategy

Literature searches were conducted in Embase (accessed through the Ovid interface, 1974 onwards), Medline (accessed through Ovid, 1946 onwards), and PsycINFO (accessed through Ovid, 1806 onwards) on 31^st^ October 2019, with no limits imposed on the searches. The search strategy used terms for ‘multiple sclerosis’ ‘functional connectivity’ and ‘cognition’ and was tailored for each database to use both controlled terms where available and uncontrolled keywords in order to capture any synonym, abbreviation and related term of the keywords of interest. The searches were repeated on 22^nd^ October 2020 to capture any studies published since the original searches. The same search strategy was used, but limits were added to capture only results which had been added or updated in the period 1^st^ November 2019 – 22^nd^ October 2020. The full search strategy used in each database is available in Supplementary Table 1.

### Study eligibility and selection

Records returned by each search were imported into the Mendeley reference management software v 1.19.4, and duplicates were removed using the tool’s de-duplication function. Titles and abstracts were then manually screened by two independent reviewers (DJ and RS). Full text publications were obtained for all papers chosen for full text review by one or both reviewers and assessed for inclusion in the review against pre-defined eligibility criteria. Any disagreements about study inclusion were resolved through discussion and reasons for study exclusion were recorded. This process was then repeated for the search conducted on 22^nd^ October 2020. The results at each stage, for the combined two searches, are presented in Figure 2.

**Figure 2.**
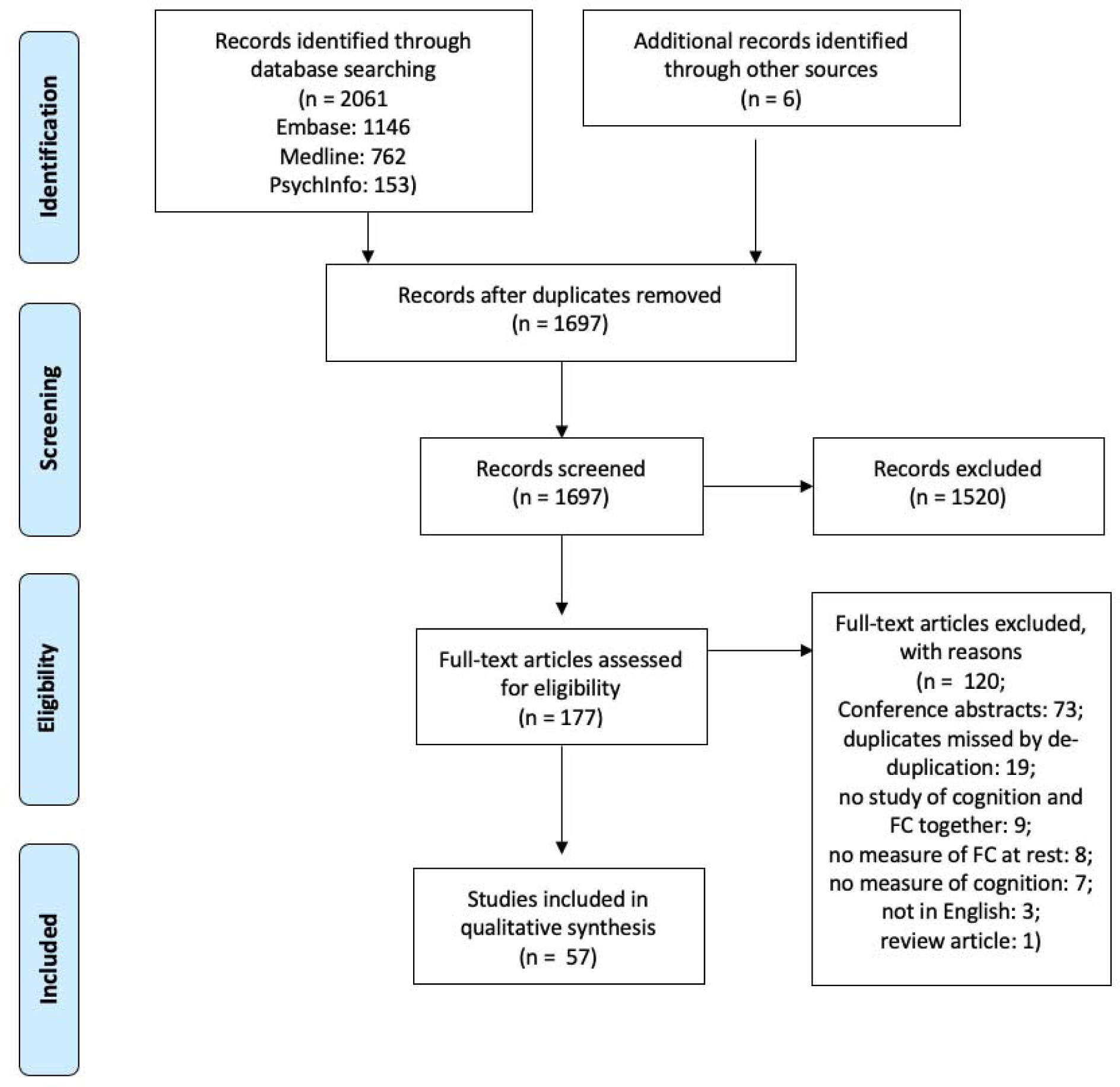
Flow diagram showing identification, screening and selection of records. Figure 1 outlines combined database searches conducted on the 31^st^ October 2019 and on the 22^nd^ October 2020 using the PRISMA protocol for studies of rs-fMRI and cognitive function in MS. Template from Moher *et al.* (2015)

Eligibility criteria were: original peer-reviewed research studies reporting on cognitive function and FC metrics derived from rs-fMRI in adult MS patients. Review articles, book chapters and conference abstracts were excluded, as were any original research studies in a paediatric population, on diseases other than MS, studies which had not measured cognitive function and/or functional connectivity, studies focusing on cognitive rehabilitation, studies which had assessed social cognition only, and any articles which were not available in English.

### Data collection and synthesis

Data extraction was performed by DJ and RS and the following data items were recorded: 1) study characteristics (authors, year of publication, journal); 2) aims of the study; 3) Participant details (MS subtype, control group, sample size, disease duration of MS sample, Expanded Disability Status Scale (EDSS) score of MS sample); 4) MR methodology (scanner field strength, MR metrics); 5) FC analysis (data pre-processing, method for analysis, whether analysis was global or regional [and if so, which regions], use of covariates); 6) cognitive testing (cognitive test(s) used, definition of cognitive impairment, number of cognitively impaired/preserved patients if applicable); 7) results from FC analysis and from other MR metrics).

To understand whether there might be a link between methodological aspects and FC results, we examined whether a particular feature was commonly present in studies that report links between worse cognition and either high or low FC. The features we examined were the MS subgroup studied, the average disease duration of patient samples, the rs-fMRI analysis method and the brain region or resting state network (RSN) investigated. Because the studies included were too heterogeneous for a meta-analysis, data synthesis was done by tallying the number of studies sharing a specific methodological feature or FC result.

### Assessment of study quality

A quality assessment approach was chosen over a risk of bias tool because most articles for inclusion in this review were expected to be cross-sectional. The AXIS tool was designed for cross-sectional studies across a range of scientific disciplines (46) and was therefore selected to judge the quality of the evidence included in the review. The AXIS tool is a 20 item checklist which asks ‘yes/no’ questions about important elements of a study. Three of the 20 items in the tool were not relevant for the studies selected for this review, as they refer to responding to an intervention, so quality assessment was based on the remaining 17 items. The items of the AXIS tool are not scored, but instead recorded in a similar way to the Cochrane risk of bias tool (47), allowing review authors to make an overall assessment of the quality of the study based on the presence or absence of reporting of the items covered by the tool.

## Results

### Study selection and quality assessment

The systematic review process is outlined in Figure 2. The database searches yielded 2061 results, and in addition 6 were identified from other sources. After removal of duplicates 1697 remained, which were screened for eligibility until 177 remained for full-text assessment. At this point 120 records were excluded, most of which were conference abstracts (see Figure 1 for reasons for exclusion). Fifty-seven studies met eligibility criteria and were included in the review. These studies are summarised in Table 1. All studies were of high quality, as measured by the AXIS tool (46). Eighteen studies did not include clear details of where participants were recruited from for the study, and very few studies (5/57) had a justification for the sample size used.

**Table 1.**
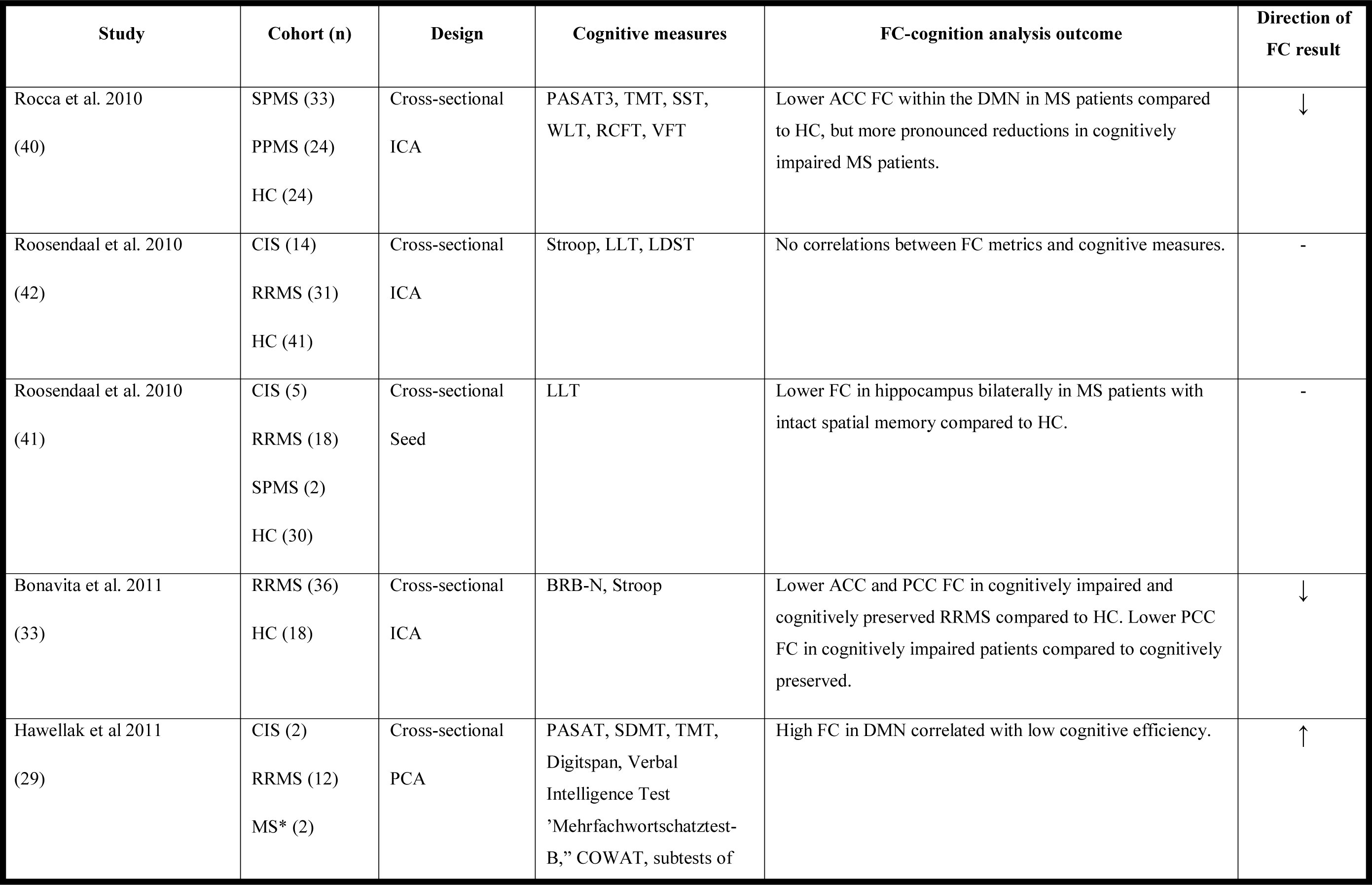
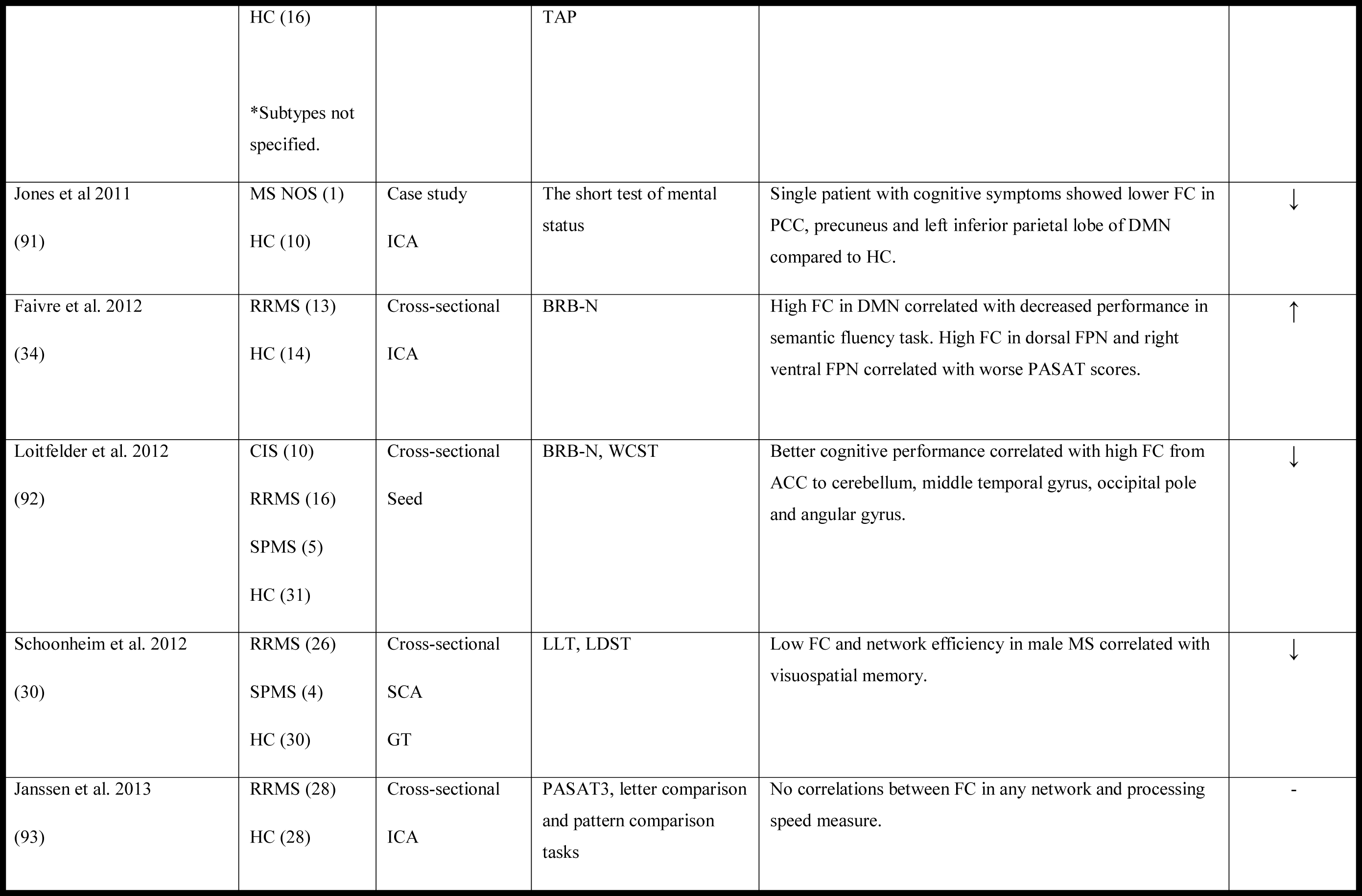
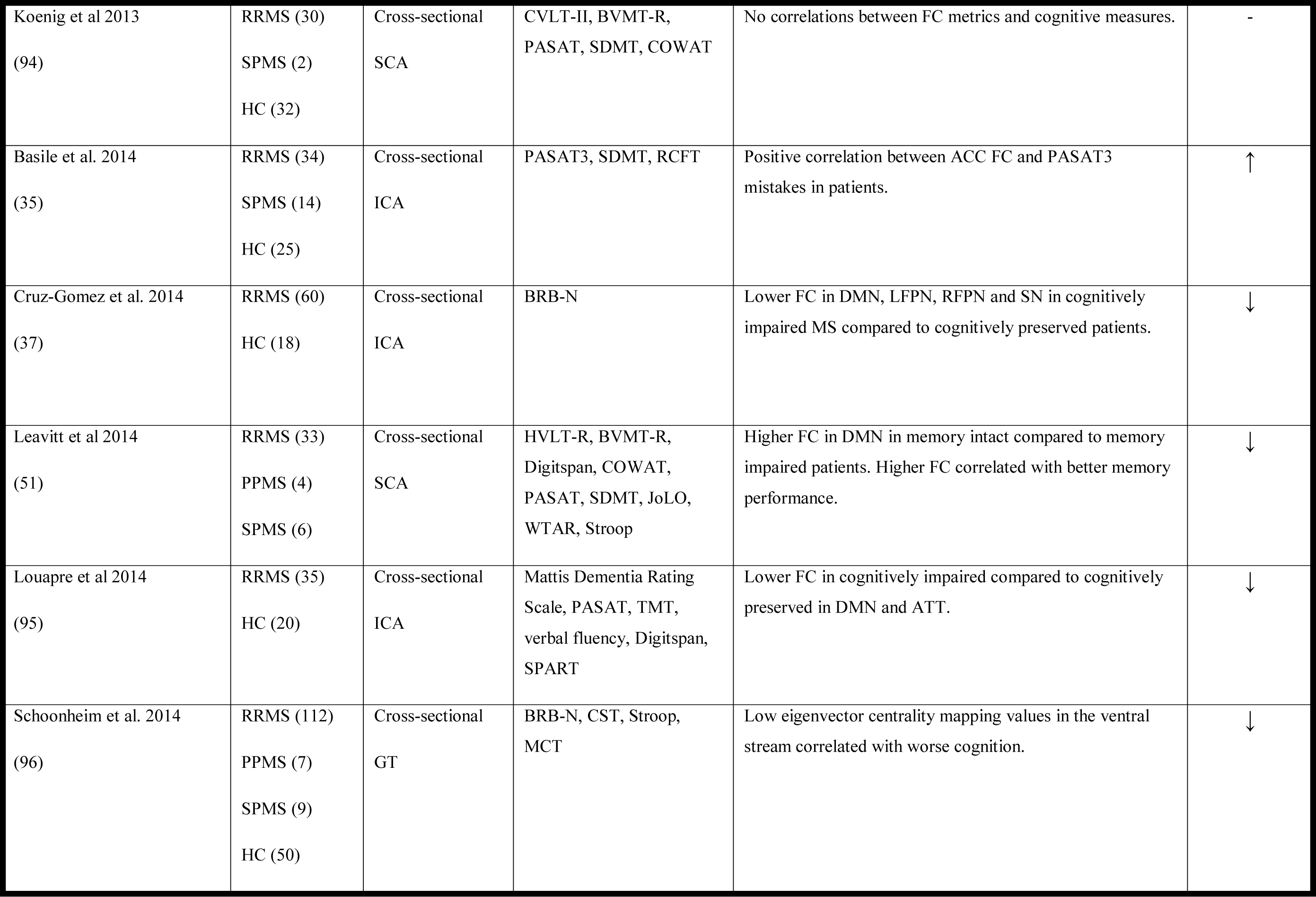
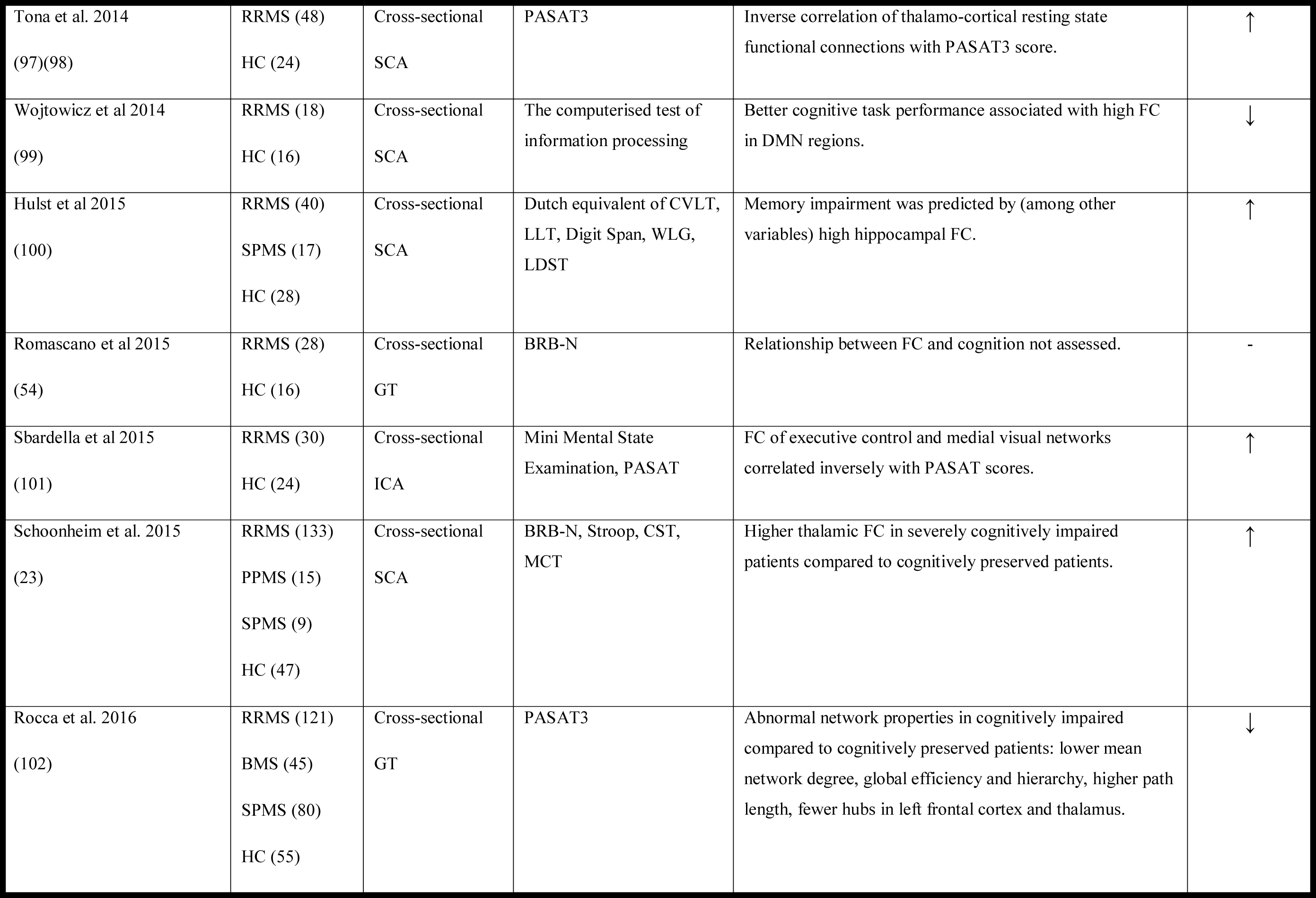
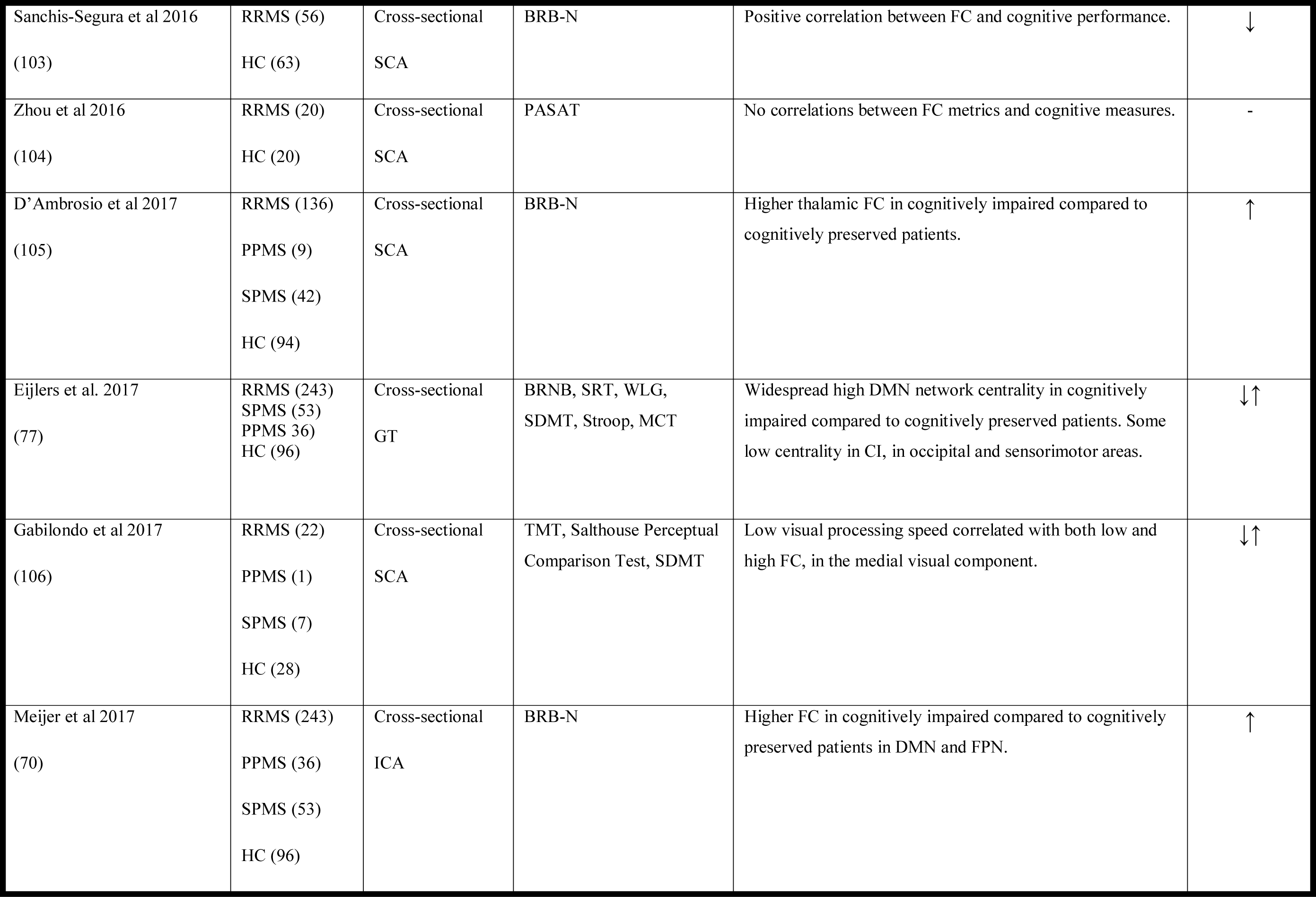
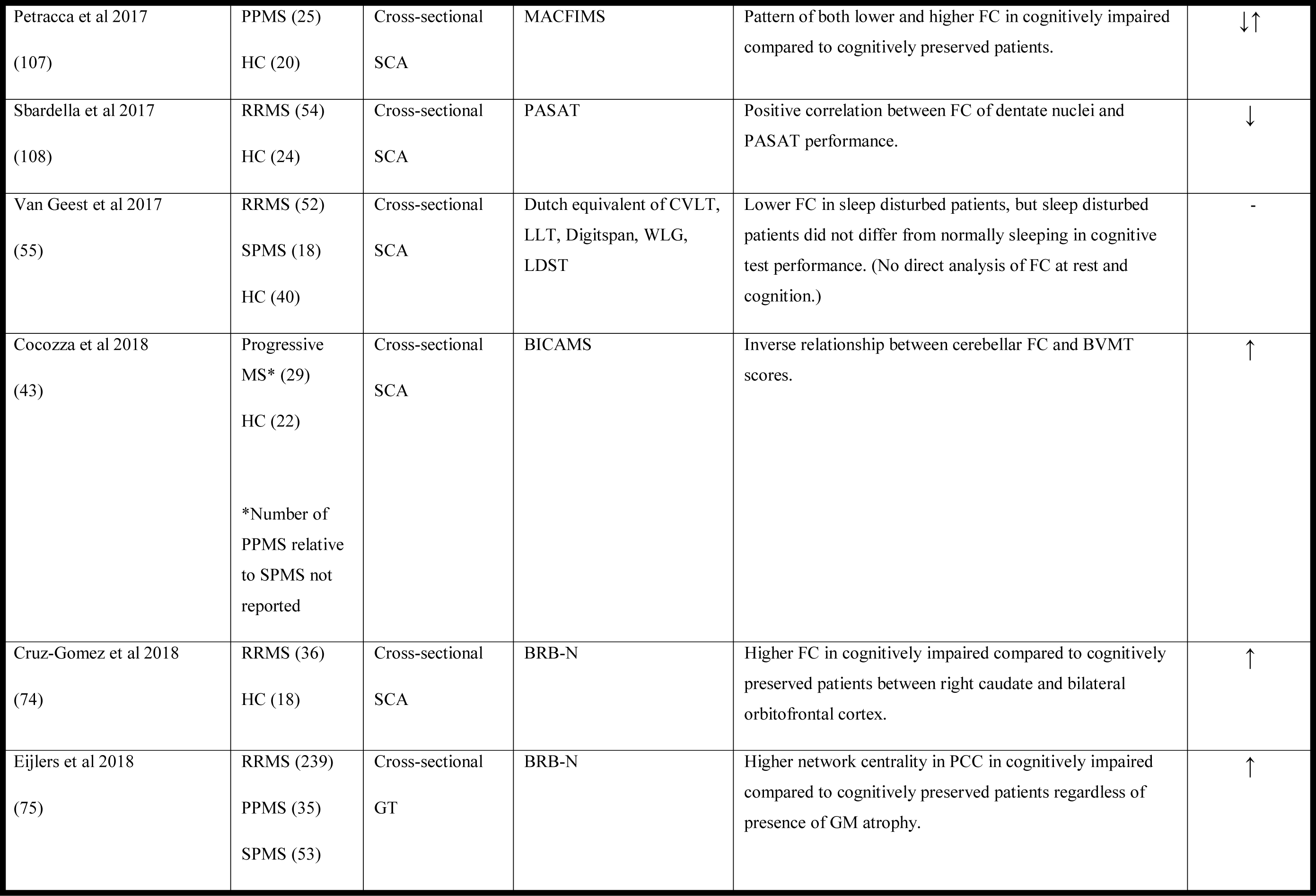
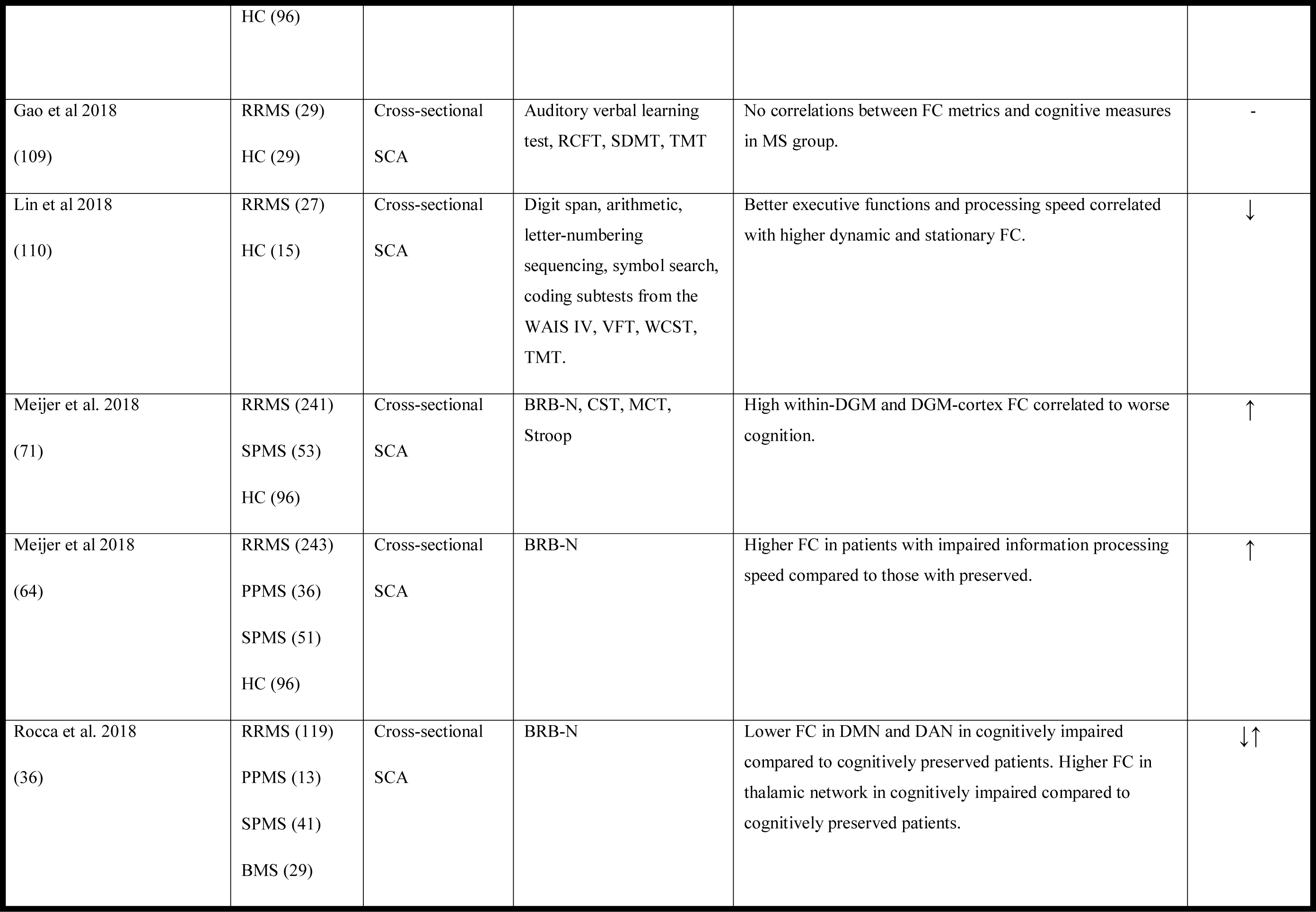
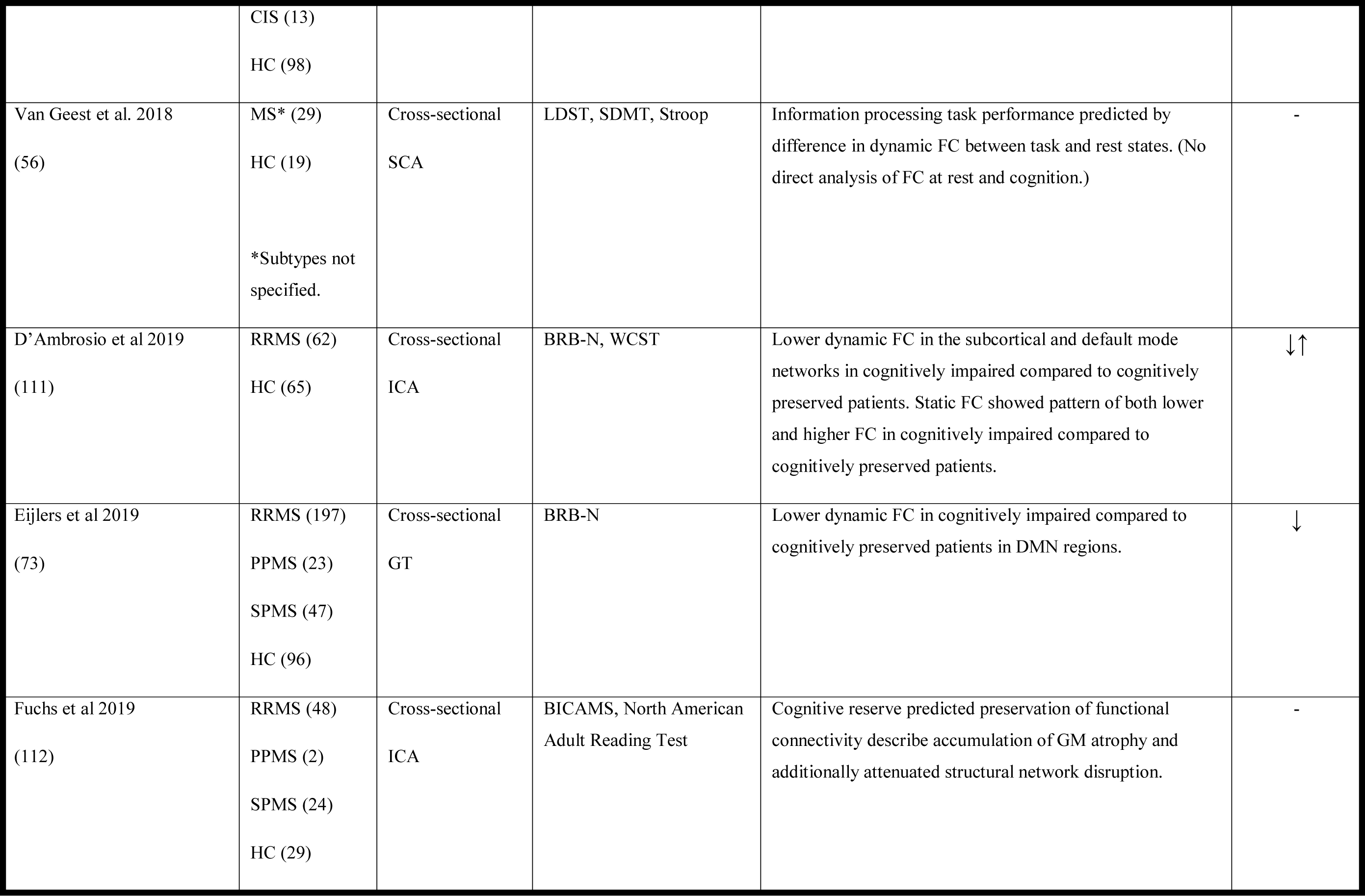
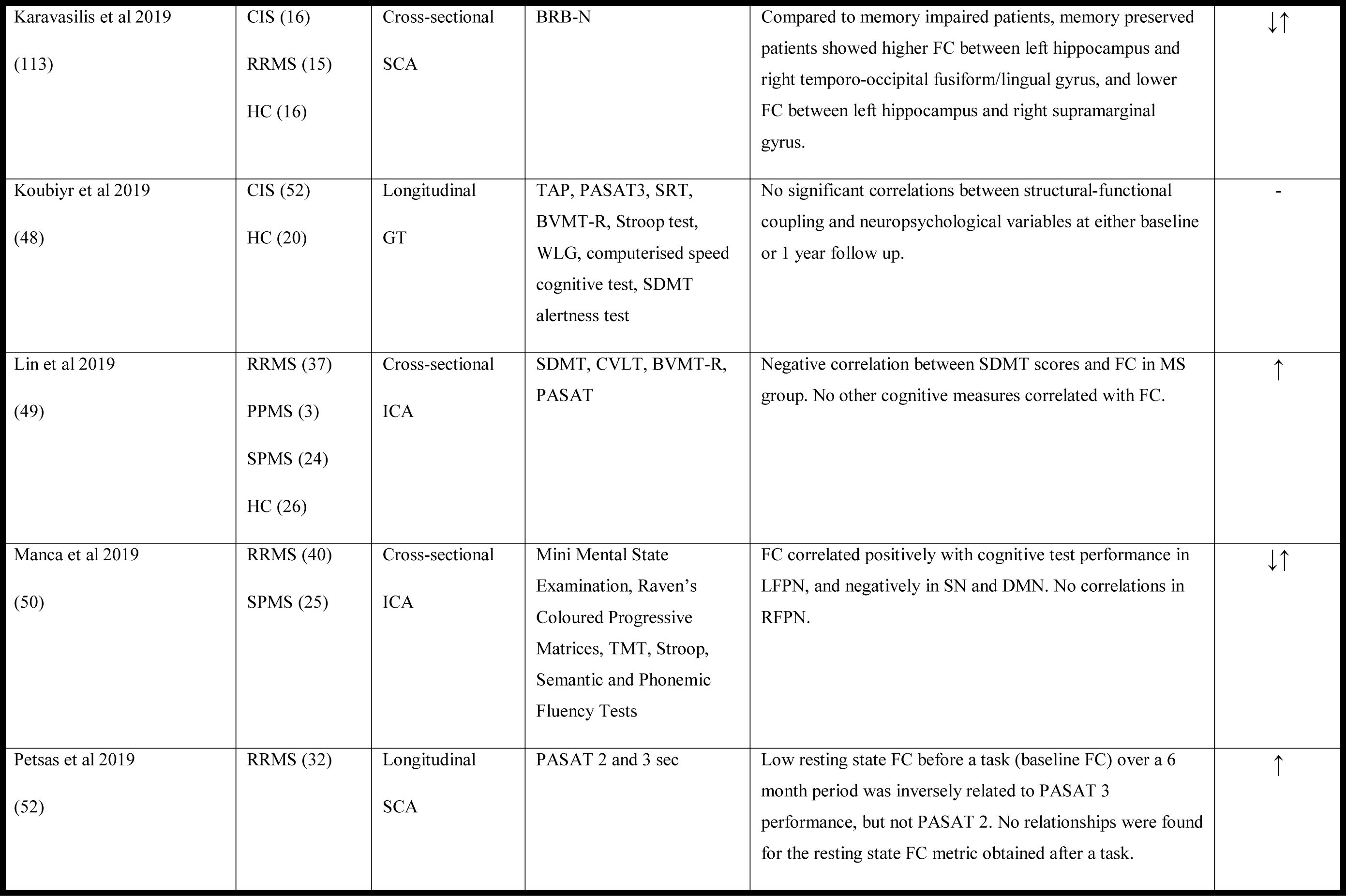
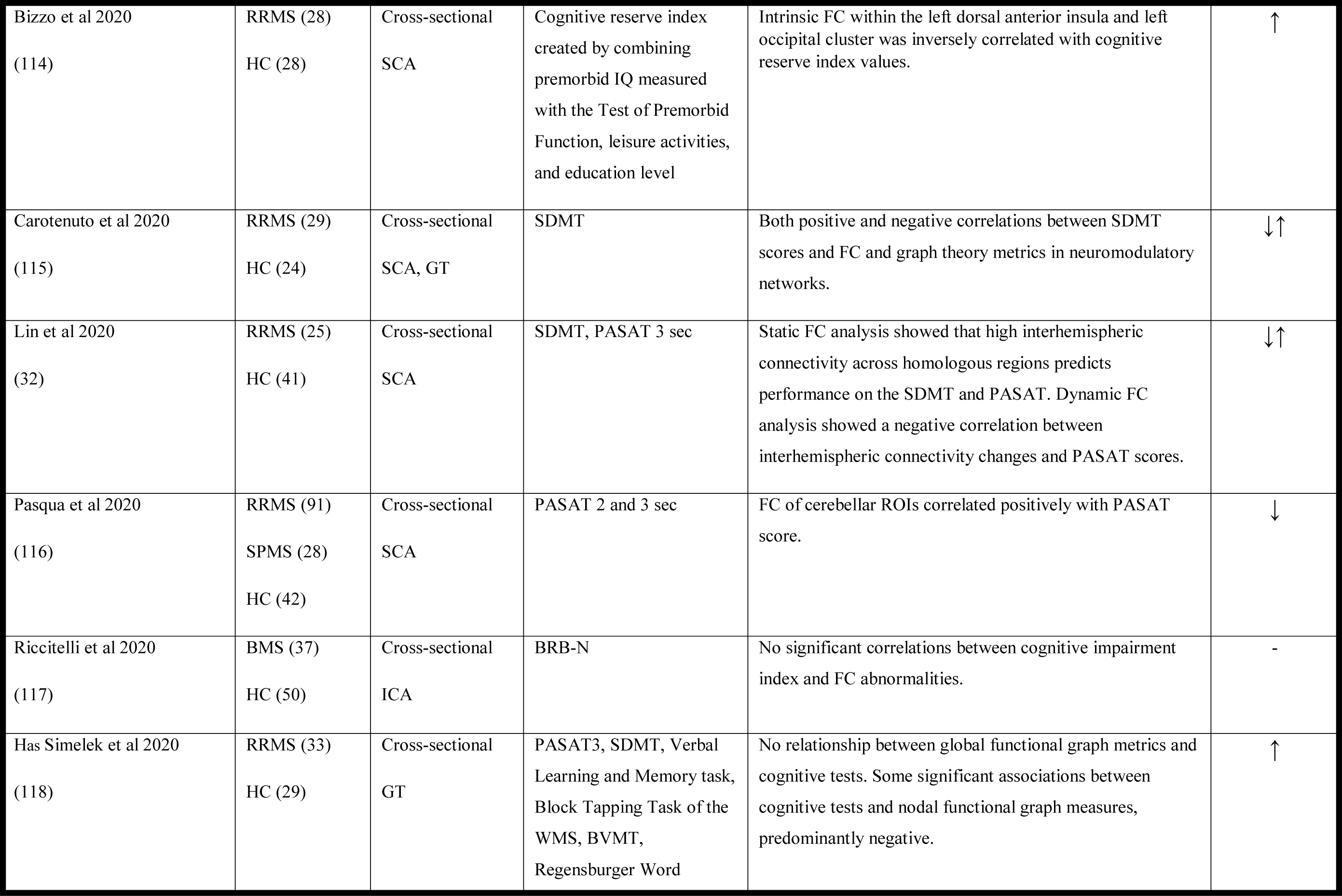
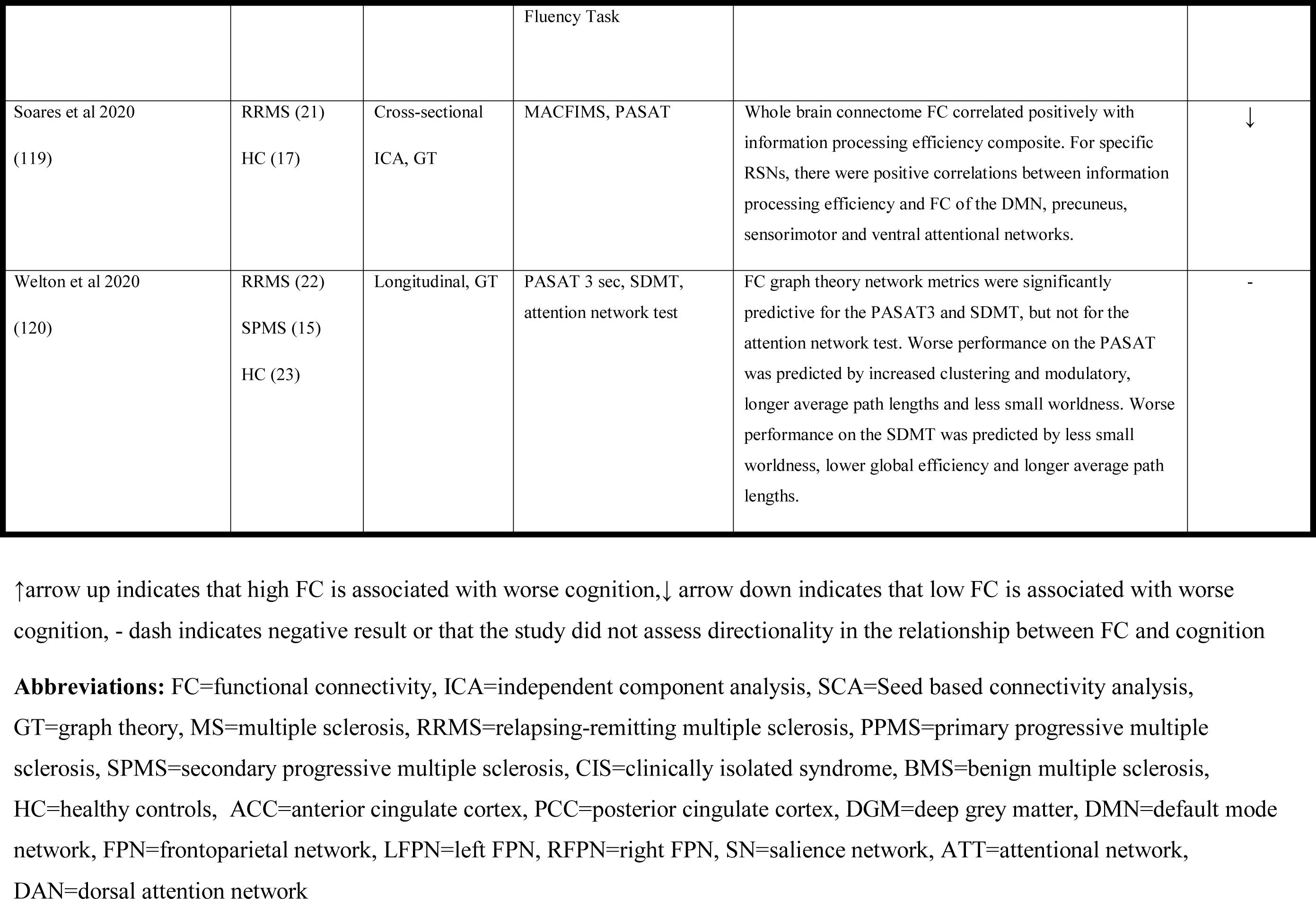
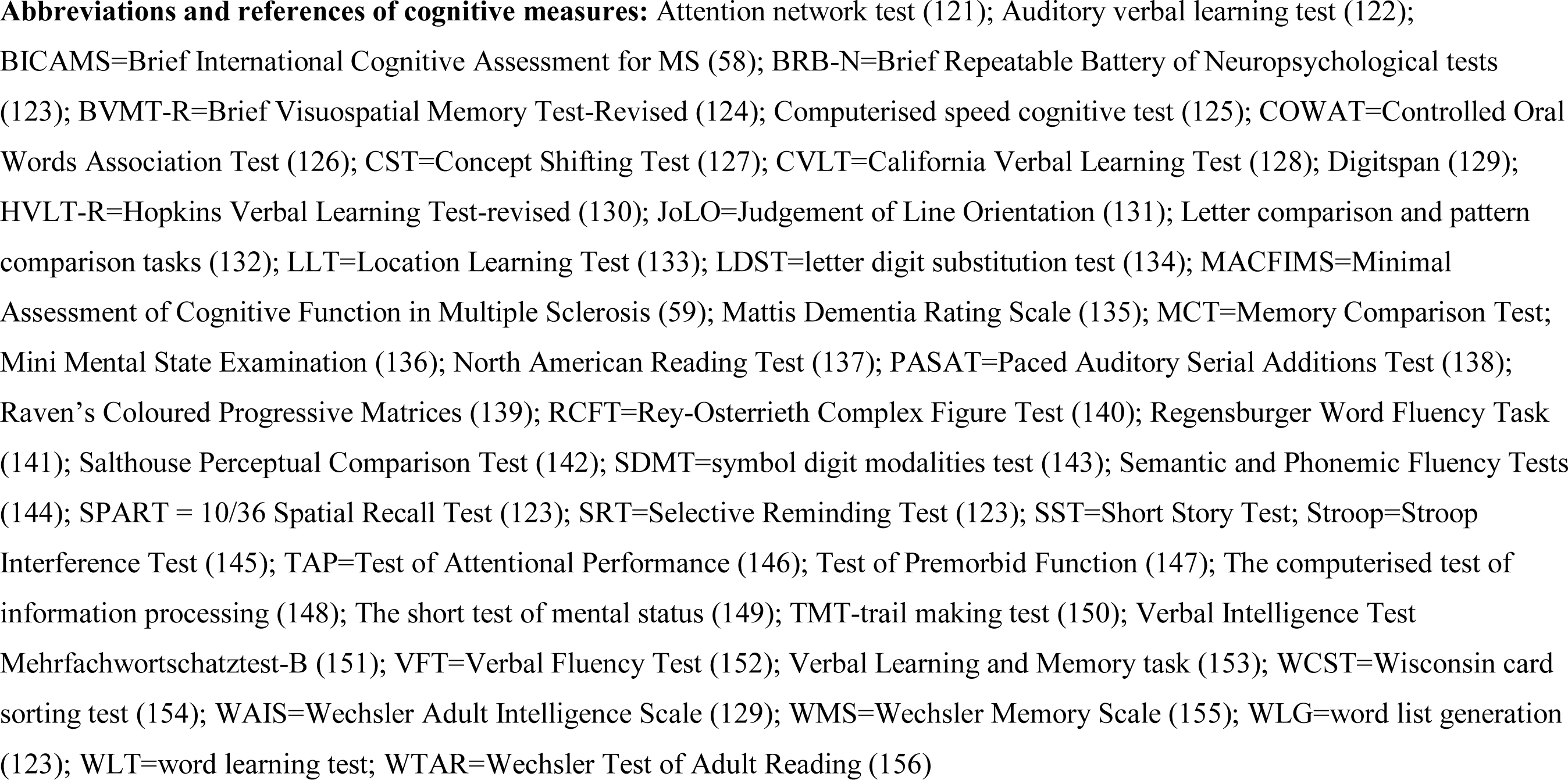
Study characteristics, cognitive assessment and relationship between cognition and functional connectivity.

### Participant characteristics

The studies that were included differed in the clinical and demographic details of the MS samples used. The majority of studies used a mixed sample of different MS phenotypes (29/57 studies), and slightly over a third used a sample of relapsing-remitting MS (RRMS) patients only (22/57 studies). The remaining six studies used either a primary progressive MS (PPMS) sample (1/57), CIS sample (2/57), a benign MS (BMS) sample (1/57) or did not specify the MS subtype (2/57). See Table 1 for details on the cohort of each study.

The average disease duration ranged from as little as 4.2 months (48) to 21.9 years (49) from either time from first symptom or from diagnosis, and median EDSS ranging from 1 (34, 48) to 6.5 (50).

Most studies (54/57) used healthy volunteers as a control group. In one study normative data from age-matched healthy controls was used for neuropsychological assessments, but no control group was used for comparisons of MRI metrics (50). In one study no control group was specified (51), and in one longitudinal study no control group was used (52). Out of the studies using healthy controls, many did not report matching groups on any demographic variables (18/54) while some reported matching groups but not on which variables (3/54) and one reported not matching the groups. Of the studies reporting the variables groups were matched on, most were on age and sex (15/54), followed by age, sex and education (10/54), age only (2/54), sex only (2/54) or age, sex, education and premorbid IQ (1/54). In this review we have interpreted the words ‘sex’ and ‘gender’ to both refer to sex, given that MS is a disease characterised by sex differences in prevalence (2, 53).

### Neuropsychological assessment

Most studies (34/57) looked at relationships between cognitive test performance and MR metrics through correlations or regressions, and 19 studies examined group differences in MR metrics between patients who met criteria for cognitive impairment and those who did not. Of the remaining four studies, one looked at FC only in MS patients with intact spatial memory (41), and three did not directly assess the relationship between cognition and FC. Despite this, they were included in the review for the following reasons: the authors of one study expressed intentions to correlate FC measures with clinical measures, but did not because the FC measure did not show any abnormalities in MS patients (54); two studies indirectly explored the relationship between FC and cognition and did not meet any exclusion criteria (55, 56).

To assess cognitive function most studies used either the Brief Repeatable Battery of Neuropsychological tests (BRB-N), which has been validated for use in MS (57), alone or in combination with other tests (20/57), or a collection of individual tests (14/57). The remaining studies used either another cognitive battery; Brief International Cognitive Assessment for MS (BICAMS) n=2 (58), Minimal Assessment of Cognitive Function in MS (MACFIMS) n=2 (59), Mini Mental State examination n=1), or a single test; Paced Auditory Serial Addition Test (PASAT) n=6, Symbol Digit Modalities Test (SDMT) n=1, Location Learning Test n=1, Short test of mental status n=1, the computerised test of information processing n=1) or a cognitive reserve index (n=1). The specific battery or tests used by each study are summarised in Table 1.

Within the 19 studies that split the MS sample into cognitively impaired and cognitively preserved sub-samples, there were 12 different definitions of cognitive impairment. Some definitions are likely guided by the test(s) used to assess cognition, but even amongst studies using the BRB-N, there were five different definitions of cognitive impairment. These include: ≥1.5 SD below normative values on ≥1 test (n=1); ≥1.5 SD below controls scores on ≥2 tests (n=5, but note that four used this definition of a mildly cognitively impaired group), ≥2 SD below normative values on ≥1 test (n=1); ≥2 SD below normative values on ≥2 tests (n=9); performance in the 5^th^ percentile of scores on either the Selective Reminding Test or Spatial Recall Test compared to normative data (n=1).

### Functional connectivity analysis

Half of all studies (28/57) used a seed-based connectivity analysis (SCA) method for assessing FC. In this category we have included studies which used one or a few specific regions of interests (ROIs; regional SCA) or divided the whole brain into ROIs and created a connectivity matrix (global SCA). The second most common method was independent component analysis (ICA) (14/57), and the remaining studies either calculated graph theory metrics (7/45), used a principal component analysis (1/45) or used a combination of SCA and graph theory (1/45) or ICA and graph theory (1/45). See Table 1 for the design and rs-fMRI analysis method of each study.

A wide range of regions and RSNs were investigated, either as a priori defined areas of interest, or as patterns emerging from a data-driven analysis, of which the most common were the DMN (21/57), thalamus and thalamic networks (9/57), the fronto-parietal network (FPN), including the right, left, dorsal and ventral FPNs (7/57). Other RSNs and regions investigated include the attentional network including left, right, dorsal, ventral variants, the salience network, the executive network, the working memory network, the motor network, the sensorimotor network, the visual processing network, the auditory network, the auditory and language processing network, visual processing networks, including medial and lateral variants, the cerebellar network, the medial prefrontal cortex, anterior cingulate cortex, posterior cingulate cortex, precuneus, basal ganglia, hippocampus and cerebellum. Ten studies conducted a whole-brain analysis and did not report regional FC changes. See Supplementary Table 2 for an overview of study results by regions investigated.

### Functional connectivity results

The main result of the relationship between FC and cognition of each study is summarised in Table 1 and Figure 3. Overall, 18 studies found worse cognition to be linked with high FC and 17 found it to be associated with low FC. Nine studies found patterns of both high and low FC to be associated with cognitive impairment, in different regions or for different MR metrics, and seven studies found no significant relationship between cognitive and FC measures. Six studies had a methodology which didn’t measure the direction of FC change in relation to cognitive impairment.

**Figure 3.**
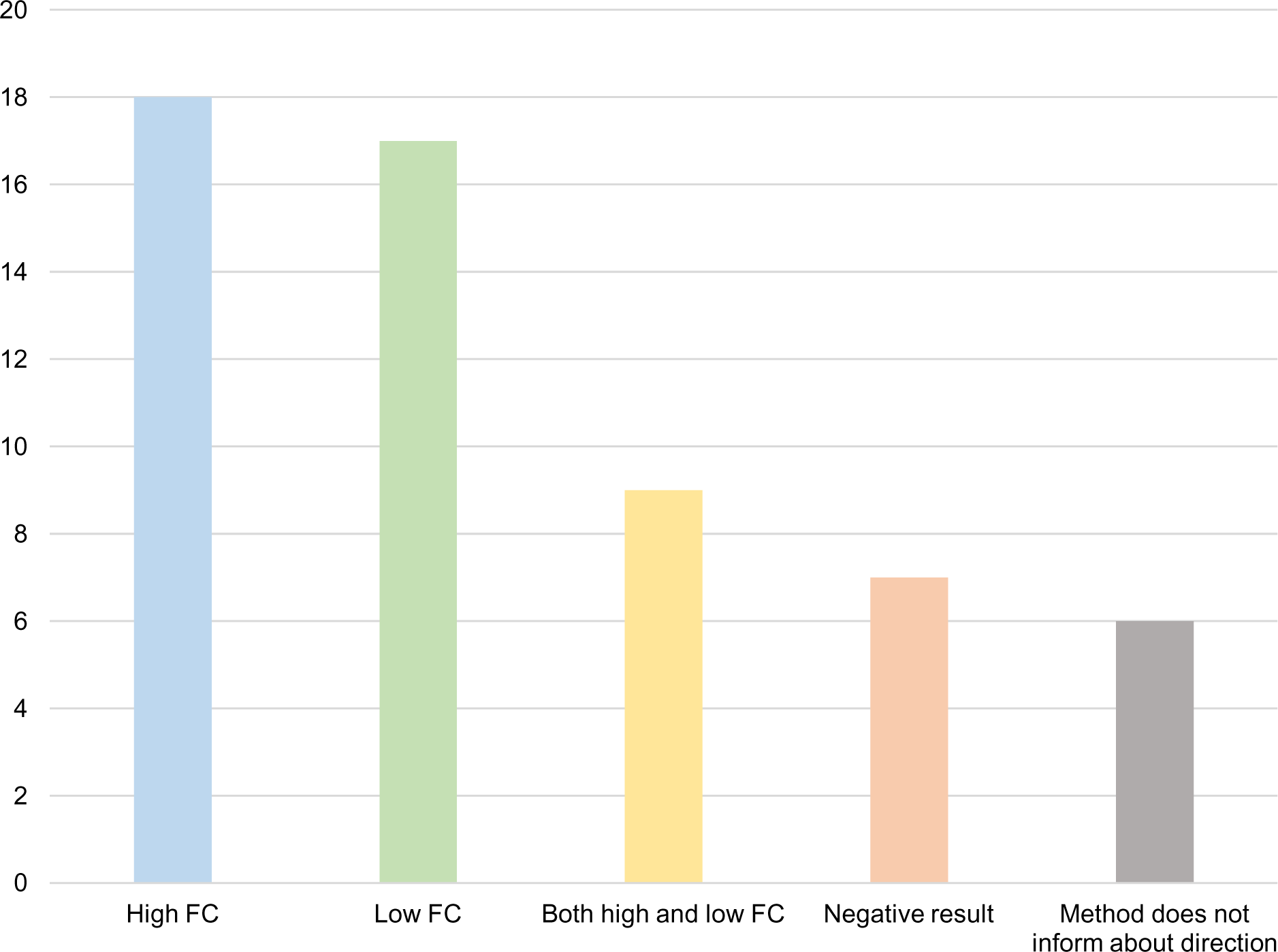
Number of studies reporting an association between poor cognitive test performance and high or low functional connectivity. Eighteen studies reported worse cognition to be associated with high functional connectivity (FC), seventeen with low FC and nine studies with both high and low FC. Seven studies did not find a link between FC abnormalities and cognitive function. Six studies used a method that does not provide information about directional changes in FC in relation to cognitive test performance. Abbreviations: FC = functional connectivity.

When grouping studies based on methodological and clinical features to assess whether one direction of FC change associated with worse cognition is more commonly seen in studies with that feature, we found no trend to suggest that one FC direction change associated with worse cognition is more commonly seen in studies using a specific method or studying a specific type of sample. This includes grouping studies based on the RSN or network assessed. Of the 21 studies measuring FC in the DMN, 10 found worse cognition to be associated with low FC, 6 with high FC, 1 with both high and low FC, 3 obtained a negative result, and one study did not test the relationship directly, see Supplementary Table 2.

We ordered studies by the average reported disease duration of the sample used, to see if patterns of FC changes differ from early to late in the disease and found no such trend, see Figure 4 and Supplementary Table 3.

**Figure 4.**
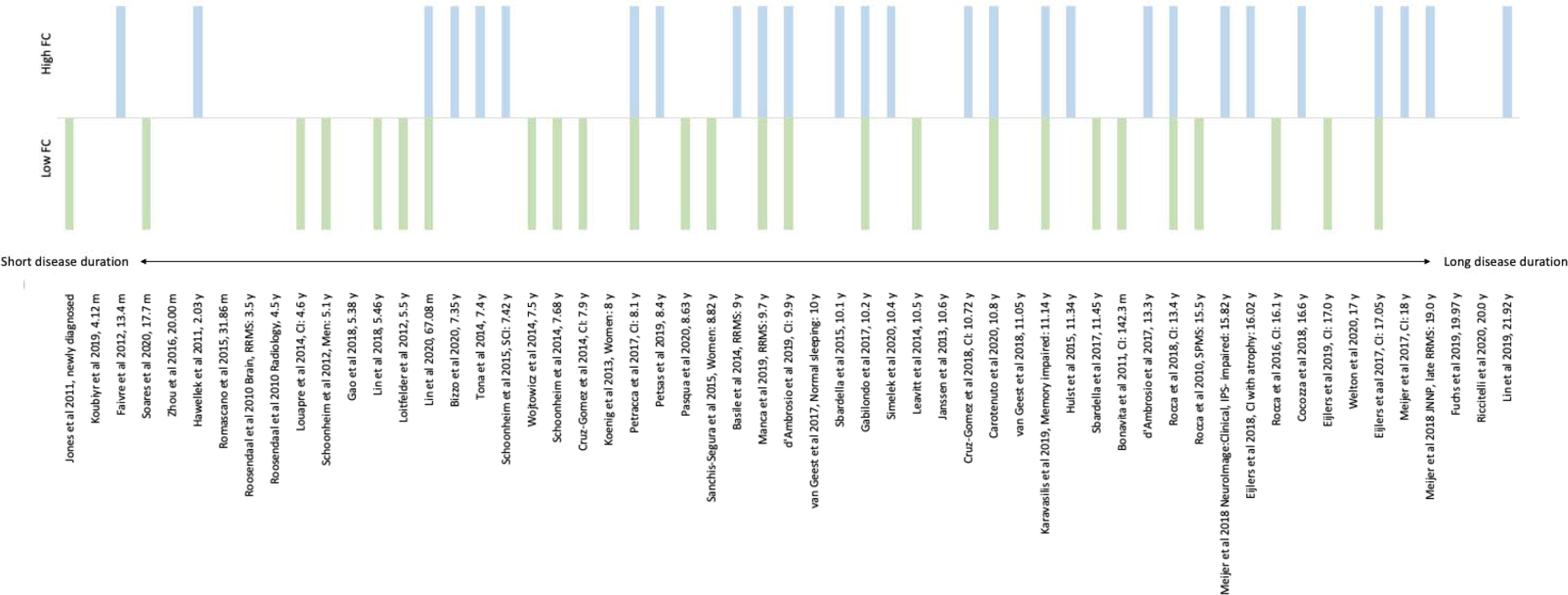
Direction of functional connectivity abnormalities sorted by average disease duration. Direction of functional connectivity abnormalities associated with worse cognition, sorted by average disease duration of the sample in each study. Because several studies used samples of mixed phenotypes and different disease durations, the following decisions were taken when ordering studies by the disease duration: 1) studies were ordered by the overall disease duration of the sample, when given; 2) studies were ordered by the disease duration of the cognitively impaired group; 3) if there were two cognitively impaired groups, studies were ordered by the disease duration of the more impaired group, or the cognitively impaired group with atrophy, in one case; 4) when the disease duration was only reported for each MS phenotype, or sex, studies were ordered by the disease duration of the larger sample; 5) for a study which had equal numbers of males and females, the study was ordered by the sex with the longer disease duration; 6) for one study that used a subset of MS patients that were matched to HC, the study was ordered by the disease duration of the matched subset. Full references are provided in Table 1.

We also considered the role of phenotype, however, most studies used either a mixed sample consisting of several phenotypes or a sample of RRMS patients only. Of the 22 studies which used a RRMS sample, eleven reported worse cognition to be associated with high FC and ten with low FC. Three studies reported a negative result and one had a study method which does not inform about the direction of FC changes. Similarly, within the mixed sample studies almost half of studies reported worse cognition to be associated with high FC (13/29) and more than half with low FC (16/29). Some studies reported both high and low FC to be associated with worse cognitive function and have therefore been counted twice. See Supplementary Figure 1 for an overview. In seven of the studies with mixed phenotype samples subgroup analyses were conducted to compare FC changes between different MS phenotypes in the sample, but only two included cognition in these analyses. One found a stronger positive correlation between FC in the DMN and errors on the PASAT in secondary progressive MS (SPMS) compared to RRMS, while another found differences between RRMS and SPMS in the spatial location of FC abnormalities that corelated with cognitive test performance.

## Discussion

In this systematic review we examined the consistency and direction of findings of studies investigating associations between rs-fMRI FC measures and cognition in MS. Overall, the studies reviewed support the notion of FC alterations associated with cognitive dysfunction in MS (Filippi and Rocca, 2013). Although most changes were related to cognitive dysfunction, the direction of FC changes varied considerably between studies and was not clearly linked to any methodological factors. There was substantial heterogeneity in clinical and rs-fMRI methodology, as has previously been noted in non-imaging cognition studies in MS (7, 9). We therefore consider ways in which the field can reflect on what has been learned to date and improve future study designs to more clearly understand the mechanisms and consequences of changes in rs-fMRI FC.

The most commonly used model for understanding FC changes in MS is the ‘network collapse’ model, which postulates three main stages (38). In the first, early stage network efficiency remains normal, at this point structural damage can be compensated by increases in local activation. This predicts early increases in FC, reflecting these compensatory processes. The second stage is where structural damage accrues to a critical point, at which compensatory processes become less effective. Finally, in the third stage structural damage exceeds the critical point with associated ‘network collapse’, and concomitant decreases in FC. Computational modelling of empirical data on FC in MS supports this model (44). Similarly, longitudinal studies demonstrate a reorganisation of structural and functional networks in early stages of MS (i.e. CIS) despite intact cognitive performance, suggesting compensatory processes are at work (48). Cross-sectional task-related fMRI studies also indicate increasing deviation from healthy control patterns of brain activation during cognitive tasks, consistent with functional reorganization, as patients progress from CIS to RRMS to secondary progressive MS (62). Together, these theories predict early adaptive reorganization of functional networks, followed by a failure of effective network organization in MS over time (see also Chard *et al.*, 2021).

In our review, when ordering studies by the average disease duration of the sample, we did not observe a trend in the direction of FC findings from early to advanced MS, as predicted by the network collapse model. We therefore consider whether the lack of fit to the model relates to the particular samples or methods of analysis employed. Many of the studies included in this review used samples of mixed phenotypes. MS phenotype has previously been reported to influence resting network FC alterations, so the inclusion of mixed MS samples could contribute to the lack of consistency in findings. However, in our review only two studies assessed the relationship between FC, phenotype and cognition, and these found both abnormally increased (64) and abnormally decreased (36) FC in patients with progressive MS. This suggests that even in specific MS subgroups, there remains considerable variability in the direction of findings. More evidence is needed in order to determine whether FC changes vary between phenotypes, and whether any model of network changes has different explanatory power for the different phenotypes. A further important consideration is the effect of disease duration and how it may mediate the relationship between FC, phenotype and cognition. Longer disease duration in RRMS is associated with FC changes in attentional, executive, and default mode networks (65). This suggests that disease duration may have an important influence on FC changes associated with cognitive impairment, possibly due to increased structural damage with longer disease duration. Those studying patients with longer disease duration (such as those with SPMS) will also have to account for age-related atrophy in these samples (66), which will be exacerbated when studying those patients with relapsing as well as progressive subtypes of MS.

We also considered whether the direction of FC change relates to definitions of cognitive impairment and choice of FC analysis. Studies of cognition in MS use a vast array of definitions of cognitive impairment (7–9), as reflected in this review. For example, of the studies using the BRB-N to assess cognitive function, most use a more conservative definition of cognitive impairment of at least 2 SDs below controls on 2 or more tests (23,36,67–73), but other, less conservative definitions are used too (37,74,75). The definition of cognitive impairment has been shown to have effects on underlying FC alterations of MS CI by the classification used (76). A few studies have compared different thresholds of cognitive impairment and found the greatest FC abnormalities in those participants meeting the more conservative thresholds (i.e. more than 2 standard deviations from controls on 2 or more tests). In contrast, less clear FC abnormalities were observed in samples performing between 1.5 and 2 SDs below controls on 2 tests (“mild cognitive impairment”) (23,70,76,77). This demonstrates the possible effect of the definition of cognitive impairment on FC findings and the arbitrary nature of these thresholds. Such findings highlight the importance of using a consistent measure of cognitive dysfunction and definition of impairment across studies. As a further challenge there is no established specific cognitive test or battery for defining cognitive dysfunction in MS despite documented phenotypic differences in impairments by test and domain (13,14,78,79). Separately, we found scant evidence to support a consistent direction of FC change in cognitively impaired patients when using model-based (e.g. seed) or data driven (e.g. ICA) approaches, or when considering specific resting state networks. Indeed the default mode network, the most commonly studied RSN across the literature, showed both increases and decreases in cognitively impaired MS patients (see supplementary Table 2).

There also needs to be a greater understanding of the mechanisms through which FC changes in MS. The ‘network collapse’ model suggests that network efficiency reduction is a function of accumulation of structural damage. In support of this, work focusing on structural connectivity in MS has found consistent evidence for structural network alterations associated with cognitive dysfunction (80–82). However, these studies have considered white matter in isolation, so conclusions about the effect of anatomical network changes including grey matter on functional connectivity cannot be drawn. In contrast, multimodal MRI studies of diffusion-weighted MRI (DWI) and rs-fMRI can assess the relationship between changes in structural and functional connectivity. Those that have been conducted support the influence of alterations in white matter linked to FC abnormalities in MS, and fit the predictions of the ‘network collapse’ model (44,83–85). Future multimodal studies using DWI and rs-fMRI can test the predictions of the ‘network collapse’ model further and to develop this or new models as needed to better characterise progression and the influence of pathology in MS brains, in order to develop clinically useful disease markers. In addition, there is evidence of physiological abnormalities in MS that are associated with cognitive dysfunction, such as cerebral hypoperfusion and sodium accumulation in the grey and white matter (86–88), and additional proton spectroscopic changes (89). Considering how these are related to network changes can help us understand the mechanisms of network abnormalities and aid in the search for a biomarker of cognitive impairment.

This systematic review provides a call to arms for the need to standardize the study of cognitive impairment in MS, but also the use of specific rs-fMRI methodology and interpretations of results. Ten years ago Fox and Greicius (2010) identified inconsistent results of FC changes across rs-fMRI studies as a barrier to the clinical applicability of this modality, and suggested a set of guidelines for rs-fMRI studies of clinical populations (90). Despite this, heterogeneity in study methodology seems to be a challenge across neurodegenerative diseases investigated by rs-fMRI, and the rs-fMRI derived FC measure is not yet suitable as a biomarker of disease (reviewed by Hohenfeld *et al.*, 2018). In this review we have shown that MS is no exception; there is considerable variability in the study of cognitive impairment in MS by rs-fMRI, both in study methods and findings. We argue that standardisation of study methods and more model-driven research would lay a clearer path towards clinical utility and the potential use as a biomarker of disease state. Despite inconsistent findings in the direction of FC abnormalities associated with worse cognitive function, most studies do show that FC alterations are a key pathological feature. A consensus on a suitable measure of cognitive dysfunction and the definition of impairment, consideration of differences between MS phenotypes and the role of aging processes, and a focus on the relationship between structural and functional connectivity changes would all be required to contribute to a better understanding of the pathology of cognitive impairment in MS.

This review is the first to systematically summarise the rs-fMRI functional connectivity literature on cognitive impairment in MS. However, there are some limitations to consider. First, rs-fMRI is not the only imaging modality for studying functional connectivity. While they were outside the scope of this review, electroencephalography and magnetoencephalography studies may offer additional insights into FC changes associated with cognitive impairment in MS. Similarly, there are other network measures that can be derived from rs-fMRI in addition to FC, such as dynamic FC and graph theory measures. At present the number of studies reporting these measures is small and so we did not consider them separately, but rather grouped them with the FC measure for the purposes of the review. Nevertheless, these metrics provide somewhat different information to the FC metric, which has not been captured in detail in this review. Finally, we did not carry out a formal statistical meta-analysis of the studies in this review. Instead, due to low numbers of homogeneous studies we were limited to tallying the number of studies with a specific feature. As studies start to become more consistent in their use of methods it will become easier to determine across the field whether the hypotheses including disease-specific effects, such as the ‘network collapse’ model, can suitably explain the patterns of associations that are observed.

In conclusion, this systematic review shows that cognitive impairment in MS is associated with both high and low FC, indicating that any network change seems related to poorer functioning. To better understand the relationship between worsened cognitive function and FC abnormalities, including directional FC changes, there must be standardisation in the field of the definition and measurement of CI, rs-fMRI methodology, and correction and allowances for MS phenotype, lesional changes, and non-MS related pathology from ageing.

## Abbreviations

BICAMS: Brief International Cognitive Assessment for Multiple Sclerosis
BMS: Benign Multiple Sclerosis
BOLD: Blood-Oxygenation-Level-Dependent
CI: Cognitively Impaired
CIS: Clinically Isolated Syndrome
CP: Cognitively Preserved
DMN: Default Mode Network
EDSS: Expanded Disability Status Scale
FC: Functional Connectivity
ICA: Independent Component Analysis
MACFIMS: Minimal Assessment of Cognitive Function in Multiple Sclerosis
MRI: Magnetic Resonance Imaging
MS: Multiple Sclerosis
PASAT: Paced Auditory Serial Addition Test
PPMS: Primary Progressive Multiple Sclerosis
PRISMA: Preferred Reporting Items for Systematic Reviews and Meta-Analysis
PROSPERO: International Prospective Register of Systematic Reviews
ROI: Region Of Interest
RRMS: Relapsing-Remitting Multiple Sclerosis
Rs-fMRI: Resting State Functional MRI
RSN: Resting State Network
SCA: Seed Based Connectivity Analysis
SDMT: Symbol Digit Modalities Test
SPMS: Secondary Progressive Multiple Sclerosis

