## Supplementary material for "A systematic review of resting state functional MRI connectivity changes and cognitive impairment in multiple sclerosis"

**Table 1**. **Search strategies for literature searches in Embase, Medline and PsychINFO. Searches were conducted on the 31^st^ October 2019 and updated on 22^nd^ October 2020.**

| Database: Embase <1974 to 2019 Week 43>  Search Strategy: | Database: Ovid MEDLINE(R) <1946 to October Week 4 2019>  Search Strategy: | Database: PsycINFO <1806 to October Week 3 2019>  Search Strategy: |
| --- | --- | --- |
| 1 multiple sclerosis/ (122811)  2 functional magnetic resonance imaging/ (74477)  3 functional neuroimaging/ (11826)  4 functional connectivity/ (12717)  5 resting state network/ (5850)  6 rsfmri.mp. (703)  7 rs fmri.mp. (2131)  8 rs-fmri.mp. (2131)  9 resting state fmri.mp. (6119)  10. resting state functional magnetic resonance imaging.mp. (3664)  11 network.mp. (488049)  12 connectivity.mp. (69254)  13 cognition/ (227236)  14 cognitive defect/ (157776)  15 cogniti*.mp. (649243)  16 symptom*.mp. (1750241)  17 impairment*.mp. (584165)  18 dysfunction*.mp. (784129)  19 decline.mp. (264441)  20 defect*.mp. (830373)  21 deficit*.mp. (347374)  22 disabilit*.mp. (317502)  23 problem*.mp. (1262696)  24 2 or 3 or 4 or 5 or 6 or 7 or 8 or 9 or 10 or 11 or 12 (575026)  25 16 or 17 or 18 or 19 or 20 or 21 or 22 or 23 (5023274)  26 15 and 25 (404197)  27 13 or 14 or 26 (516348)  28 1 and 24 and 27 (1002)    Limits added to search conducted 22^nd^ October 2020:  29 limit 28 to dd=20191101-20201022 (61)  30 limit 28 to rd=20191101-20201022 (83)  31 29 or 30 (144) | 1 Multiple Sclerosis/ (50628)  2 Magnetic Resonance Imaging/ (385453)  3 Functional Neuroimaging/ (3309)  4 rsfmri.mp. (225)  5 rs fmri.mp. (801)  6 rs-fmri.mp. (801)  7 resting state fmri.mp. (2674)  8 resting state functional magnetic resonance imaging.mp. (2035)  9 network.mp. (256415)  10 connectivity.mp. (39575)  11 Cognition/ (92646)  12 Cognition Disorders/ (63435)  13 Cognitive Dysfunction/ (13910)  14 cogniti*.mp. (355865)  15 symptom*.mp. (954357)  16 impairment*.mp. (285360)  17 dysfunction*.mp. (413051)  18 decline.mp. (169699)  19 defect*.mp. (474313)  20 deficit*.mp. (215585)  21 disabilit*.mp. (231078)  22 problem*.mp. (889159)  23 2 or 3 or 4 or 5 or 6 or 7 or 8 or 9 or 10 (647967)  24 15 or 16 or 17 or 18 or 19 or 20 or 21 or 22 (3126890)  25 14 and 24 (196009)  26 11 or 12 or 13 or 25 (263948)  27 1 and 23 and 26 (639)    Limits added to search conducted 22^nd^ October 2020:  28 limit 27 to dt=20191101-20201022 (11)  29 limit 27 to rd=20191101-20201022 (123)  30 28 or 29 (123) | 1 exp Multiple Sclerosis/ (12466)  2 exp Functional Magnetic Resonance Imaging/ (21226)  3 brain connectivity/ (4435)  4 rsfmri.mp. (188)  5 rs fmri.mp. (614)  6 rs-fmri.mp. (614)  7 resting state fmri.mp. (2198)  8 resting state functional magnetic resonance imaging.mp. (1687)  9 network.mp. (91298)  10 connectivity.mp. (24043)  11 exp Cognition/ (36363)  12 exp Cognitive Impairment/ (35048)  13 cogniti*.mp. (576040)  14 symptom*.mp. (352302)  15 impairment*.mp. (146444)  16 dysfunction*.mp. (82048)  17 decline.mp. (47882)  18 defect*.mp. (26404)  19 deficit*.mp. (140889)  20 disabilit*.mp. (144151)  21 problem*.mp. (564617)  22 2 or 3 or 4 or 5 or 6 or 7 or 8 or 9 or 10 (118304)  23 14 or 15 or 16 or 17 or 18 or 19 or 20 or 21 (1204462)  24 13 and 23 (234956)  25 11 or 12 or 24 (259865)  26 1 and 22 and 25 (132)    Limits added to search conducted 22^nd^ October 2020:  27 limit 26 to ch=20191101-20201022 (11)  28 limit 26 to up=20191101-20201022 (11)  29 27 or 28 (21) |

**Table 2.** **Study findings sorted by the brain region or network investigated**

| **Study*/Brain region or network** | Wholebrain analysis, paper does not specify regional changes | mPFC | ACC | PCC | Precuneus | DMN | FPN incl left, right, dorsal, ventral | Attentional network incl dorsal, ventral, right, left | SN | EC | Working memory network | Motor network | Sensorimotor | Somatomotor network | Visual processing networks, incl medial and lateral | Auditory and language processing network | Auditory network | Hippocampus | Thalamus incl thalamic network | Basal ganglia | Cerebellum | Cerebellar network | Other | Other, additional (if other pattern reported) |
| --- | --- | --- | --- | --- | --- | --- | --- | --- | --- | --- | --- | --- | --- | --- | --- | --- | --- | --- | --- | --- | --- | --- | --- | --- |
| Rocca 2010 |  | ↓ | ↓ |  |  | ↓ |  |  |  |  |  |  |  |  |  |  |  |  |  |  |  |  |  |  |
| Roosendaal et al 2010  Brain |  |  |  |  |  | - | - | - |  | - |  |  | - |  | - | - |  |  |  |  |  |  |  |  |
| Roosendaal et al 2010  Radiology |  |  |  |  |  |  |  |  |  |  |  |  |  |  |  |  |  |  |  |  |  |  |  |  |
| Bonavita et al 2011 MSJ |  |  | ↓ | ↓ |  | ↓ |  |  |  |  |  |  |  |  |  |  |  |  |  |  |  |  |  |  |
| Hawellek et al 2011 PNAS |  |  |  |  |  | ↑ |  |  |  |  |  |  |  |  |  |  |  |  |  |  |  |  |  |  |
| Jones et al 2011 Arch Neurol |  |  |  | ↓ | ↓ | ↓ |  |  |  |  |  |  |  |  |  |  |  |  |  |  |  |  |  |  |
| Faivre et al 2012 MSJ |  |  |  |  |  | ↑ | ↑ |  |  |  |  |  |  |  |  |  |  |  |  |  |  |  |  |  |
| Loitfelder et al 2012 PLoS ONE |  |  | ↓ |  |  |  |  |  |  |  |  |  |  |  |  |  |  |  |  |  |  |  |  |  |
| Schoonheim et al 2012 MSJ | ↓ |  |  |  |  |  |  |  |  |  |  |  |  |  |  |  |  |  |  |  |  |  |  |  |
| Janssen et al 2013 Neurpsychologia |  |  |  |  |  | - | - |  |  | - |  | - |  |  | - |  | - |  |  |  |  | - |  |  |
| Koenig et al 2013 American Journal of Neuroradiology |  |  |  | - |  |  |  |  |  |  |  |  |  |  |  |  |  |  |  |  |  |  |  |  |
| Basile et al 2014 MSJ |  |  | ↑ |  |  | ↑ |  |  |  |  |  |  |  |  |  |  |  |  |  |  |  |  |  |  |
| Cruz-Gomez et al 2014 MSJ |  |  |  |  |  | ↓ | ↓ |  | ↓ |  |  |  |  |  |  |  |  |  |  |  |  |  |  |  |
| Leavitt et al 2014 Journal of the International Neuropsychological Society |  |  |  |  |  | ↓ |  |  |  |  |  |  |  |  |  |  |  |  |  |  |  |  |  |  |
| Louapre et al 2014 HBM |  |  |  |  |  | ↓ |  | ↓ |  |  |  | - |  |  |  |  |  |  |  |  |  |  |  |  |
| Schoonheim et al 2014 MSJ |  |  |  |  |  |  |  |  |  |  |  |  |  |  |  |  |  |  |  |  |  |  | Ventral stream ↓ |  |
| Tona et al 2014 Radiology |  |  |  |  |  |  |  |  |  |  |  |  |  |  |  |  |  |  | ↑ |  |  |  |  |  |
| Wojtowicz et al 2014 MSJ |  |  |  |  |  | ↓ |  |  |  |  |  |  |  |  |  |  |  |  |  |  |  |  |  |  |
| Hulst et al 2015 MSJ |  |  |  |  |  |  |  |  |  |  |  |  |  |  |  |  |  | ↑ |  |  |  |  |  |  |
| Romascano et al 2015 HBM |  |  |  |  |  |  |  |  |  |  |  |  |  |  |  |  |  |  |  |  | - |  |  |  |
| Sbardella et al 2015 MSJ |  |  |  |  |  |  |  |  |  | ↑ |  |  |  |  | ↑ |  |  |  |  |  |  |  |  |  |
| Schoonheim et al 2015 Neurology |  |  |  |  |  |  |  |  |  |  |  |  |  |  |  |  |  |  | ↑ |  |  |  |  |  |
| Rocca et al 2016 Brain Struct Func |  |  | ↓ |  | ↓ |  |  |  |  |  |  |  |  |  |  |  |  |  | ↓ |  | ↓ |  | Left frontal cortex, left superior frontal gyrus ↓ |  |
| Sanchis-Segura et al 2015 Neuroscience Letters |  |  |  |  |  |  |  |  |  |  |  |  |  |  |  |  |  |  |  |  |  |  | Three intra-hemispheric pathways: right olfactory cortex to right amygdala, right middle temporal pole to right inferior frontal gyrus, left parahippocampal gyrus to left inferior frontal gyrus ↑ |  |
| Zhou et al 2016 Frontiers in Human Neuroscience |  |  |  |  |  |  |  |  |  |  |  |  |  |  |  |  |  |  | - |  |  |  |  |  |
| d'Ambrosio et al 2017 HBM |  |  |  |  |  |  |  |  |  |  |  |  |  |  |  |  |  |  | ↑ |  |  |  |  |  |
| Eijlers et al 2017 Neurology |  |  |  | ↑ | ↑ | ↑ |  |  |  |  |  |  |  |  |  |  |  |  |  |  |  |  | Angular gyrus, middle parts of superior and middle frontal gyri ↑ | Right middle temporal gyrus ↓ |
| Gabilondo et al 2017 MSJ |  |  |  |  |  |  |  |  |  |  |  |  |  |  |  |  |  |  |  |  |  |  | Medial visual component ↓↑ |  |
| Meijer et al 2017 Neurology |  |  |  |  |  | ↑ | ↑ |  |  |  |  |  |  |  |  |  |  |  |  |  |  |  |  |  |
| Petracca et al 2017 Scientific Reports |  |  |  |  |  |  |  | ↓↑ |  | ↓ |  |  |  |  |  |  |  |  |  |  |  |  |  |  |
| Sbardella et al 2017 MSJ |  |  |  |  |  |  |  |  |  |  |  |  |  |  |  |  |  |  |  |  | ↓ |  |  |  |
| van Geest et al 2017 Journal of Neurology |  |  |  |  |  |  |  |  |  |  |  |  |  |  |  |  |  |  |  |  |  |  |  |  |
| Cocozza et al 2018 Journal of Neurology |  |  |  |  |  |  |  |  |  |  |  |  |  |  |  |  |  |  |  |  | ↑ |  |  |  |
| Cruz-Gomez et al 2018 Neuroreport |  |  |  |  |  |  |  |  |  |  |  |  |  |  |  |  |  |  |  | ↑ |  |  |  |  |
| Eijlers et al 2018 Radiology |  |  |  | ↑ |  |  |  |  |  |  |  |  |  |  |  |  |  |  |  |  |  |  | Occipital lobe ↓ |  |
| Gao et al 2018 Hippocampus |  |  |  |  |  |  |  |  |  |  |  |  |  |  |  |  |  | - |  |  |  |  |  |  |
| Lin et al 2018 HBM | ↓ |  |  |  |  |  |  |  |  |  |  |  |  |  |  |  |  |  |  |  |  |  |  |  |
| Meijer et al 2018 JNNP | ↑ |  |  |  |  |  |  |  |  |  |  |  |  |  |  |  |  |  |  |  |  |  |  |  |
| Meijer et al 2018 NeuroImage: Clinical | ↑ |  |  |  |  |  |  |  |  |  |  |  |  |  |  |  |  |  |  |  |  |  |  |  |
| Rocca et al 2018 MSJ |  |  |  |  |  | ↓ |  | ↓ |  |  |  |  | ↓ |  |  |  |  |  | ↑ |  |  | ↓ | Reward emotion network ↓ |  |
| Van Geest et al 2018 NeuroImage: Clinical |  |  |  |  |  |  |  |  |  |  |  |  |  |  |  |  |  |  |  |  |  |  |  |  |
| d'Ambrosio et al 2019 MSJ |  |  |  |  |  | ↓↑ |  |  |  |  |  |  | ↓ |  | ↓ |  | ↓ |  |  |  |  |  | Subcortical networks NOS ↓ |  |
| Eijlers et al 2019 Radiology |  |  |  |  |  | ↓ | ↓ |  |  |  |  |  |  |  |  |  |  |  | ↓ |  |  |  |  |  |
| Fuchs et al 2019 Human Brain Mapping |  |  |  |  |  |  |  |  |  |  |  |  |  |  |  |  |  |  |  |  |  |  |  |  |
| Karavasilis et al 2019 Brain Imaging and Behaviour |  |  |  |  |  |  |  |  |  |  |  |  |  |  |  |  |  | ↓↑ |  |  |  |  |  |  |
| Koubiyr et al 2019 Brain | - |  |  |  |  |  |  |  |  |  |  |  |  |  |  |  |  |  |  |  |  |  |  |  |
| Lin et al 2019 MSJ |  |  |  |  |  |  |  |  |  |  |  |  |  |  |  |  |  |  | ↑ |  |  |  |  |  |
| Manca et al 2019 Postgraduate Medicine |  |  |  |  |  | ↑ | ↓↑ |  | ↓ |  |  |  | ↑ |  |  |  |  |  |  |  |  |  |  |  |
| Petsas et al 2019 Frontiers in Neurology |  |  |  |  |  |  |  |  |  |  |  |  |  |  |  |  |  |  |  |  |  |  | Right hand's cortical representation to whole brain ↑ |  |
| Bizzo et al 2020 Journal of Neuroimaging |  |  |  |  |  |  |  |  |  |  |  |  |  |  |  |  |  |  |  |  |  |  | Dorsal anterior insula ↑ |  |
| Carotenuto et al 2020 Journal of Neurology |  |  |  |  |  |  |  |  |  |  |  |  |  |  |  |  |  |  |  |  |  |  | The serotonergic, the noradrenergic, the cholinergic, and the dopaminergic networks ↓↑ |  |
| Lin et al 2020 Frontiers in Neurology | ↓↑ |  |  |  |  |  |  |  |  |  |  |  |  |  |  |  |  |  |  |  |  |  |  |  |
| Pasqua et al 2020 MSJ |  |  |  |  |  |  |  |  |  |  |  |  |  |  |  |  |  |  |  |  | ↓ |  |  |  |
| Riccitelli et al 2020 Journal of Neurology |  |  |  |  |  | - |  |  | - | - | - |  |  |  |  |  |  |  |  |  |  |  |  |  |
| Simelek et al 2020 NeuroImage: Clinical | ↑ |  |  |  |  |  |  |  |  |  |  |  |  |  |  |  |  |  |  |  |  |  |  |  |
| Soares et al 2020 Brain Imaging and Behavior | ↓ |  |  |  | ↓ | ↓ |  | ↓ |  |  |  |  | ↓ |  |  |  |  |  |  |  |  |  |  |  |
| Welton et al 2020 Brain Connectivity |  |  |  |  |  |  |  |  |  |  |  |  |  |  |  |  |  |  |  |  |  |  |  |  |

* See main manuscript for full citations of studies.

Table presents the directional FC result for each region investigated, either as a priori defined areas of interest, or as patterns emerging from a data-driven analysis.Key: High FC is denoted by ↑ and a blue colour cell; low FC by ↓ and a green colour cell; studies demonstrating both high and low FC associated with worse cognition are denoted by ↑↓ and a yellow colour cell; studies which did not find support for a relationship between FC alterations and worse cognition are denoted by – and a pink colour cell; “something else,” denoted by a grey colour cell, refers to studies which used a methodology that does not fit into the groupings presented in this table . Abbreviations: ACC = Anterior Cingulate Cortex, DMN = Default Mode Network, EC = Executive Control network, mPFC = medial Prefrontal Cortex, NOS = Not Otherwise Specified, PCC = Posterior Cingulate Cortex, SN = Salience Network

**Table 3.** **Study findings sorted by the average disease duration of the sample**

| **Study*** | **Average disease duration**† | **Direction of FC result**‡ **associated with worse cognitive function** |
| --- | --- | --- |
| Jones et al 2011 Arch Neurol | Not specified, patient newly diagnosed | ↓ |
| Koubiyr et al 2019 Brain | 4.12 m | - |
| Faivre et al 2012 MSJ | 13.4 m | ↑ |
| Soares et al 2020 Brain Imaging and Behavior | 17.7 m | ↓ |
| Zhou et al 2016 Frontiers in Human Neuroscience | 20.00 m | - |
| Hawellek et al 2011 PNAS | 2.03 y | ↑ |
| Romascano et al 2015 HBM | 31.86 m | something else |
| Roosendaal et al 2010 Brain | CIS: 1.4 y, RRMS: 3.5 y | - |
| Roosendaal et al 2010 Radiology | 4.5 y | something else |
| Louapre et al 2014 HBM | CI: 4.6 y, CP: 4.5 y | ↓ |
| Schoonheim et al 2012 MSJ | Men: 5.1 y, Women: 4.9 y | ↓ |
| Gao et al 2018 Hippocampus | 5.38 y | - |
| Lin et al 2018 HBM | 9.85 y in overall MS group, 5.46 y in those matched to HC | ↓ |
| Loitfelder et al 2012 PLoS ONE | 5.5 y | ↓ |
| Cruz-Gomez et al 2014 MSJ | CP 5.5 y , CI 7.9 y | ↓ |
| Lin et al 2020 Frontiers in Neurology | 67.08 m | ↑↓ |
| Bizzo et al 2020 Journal of Neuroimaging | 7.35 y | ↑ |
| Tona et al 2014 Radiology | 7.4 y | ↑ |
| Schoonheim et al 2015 Neurology | CP: 7.49 y, MCI: 7.57 y, SCI: 7.42 y | ↑ |
| Wojtowicz et al 2014 MSJ | 7.5 y | ↓ |
| Schoonheim et al 2014 MSJ | 7.68 y | ↓ |
| Koenig et al 2013 American Journal of Neuroradiology | Men: 6.5 y, Women: 8 y | - |
| Petracca et al 2017 Scientific Reports | 8 y for whole group, 9.7 y for CP, 8.1 y for CI | ↑↓ |
| Petsas et al 2019 Frontiers in Neurology | 8.4 y | ↑ |
| Pasqua et al 2020 MSJ | 8.63 y | ↓ |
| Sanchis-Segura et al 2015 Neuroscience Letters | Females: 8.82 y, Males: 6.45 y | ↓ |
| Basile et al 2014 MSJ | RRMS: 9 y, SPMS: 13 y | ↑ |
| Manca et al 2019 Postgraduate Medicine | RRMS: 9.7 y, SPMS: 15.5 y | ↑↓ |
| d'Ambrosio et al 2019 MSJ | 8.2 y (7.1 y for CP, 9.9 y for CI) | ↑↓ |
| van Geest et al 2017 Journal of Neurology | Normal sleeping: 10 y, sleep disturbed: 12 y | something else |
| Sbardella et al 2015 MSJ | 10.1 y | ↑ |
| Gabilondo et al 2017 MSJ | 10.2 y | ↑↓ |
| Simelek et al 2020 NeuroImage: Clinical | 10.4 y | ↑ |
| Leavitt et al 2014 Journal of the International Neuropsychological Society | 10.5 y | ↓ |
| Janssen et al 2013 Neurpsychologia | 10.6 y | - |
| Cruz-Gomez et al 2018 Neuroreport | CP: 5.94 y, CI: 10.72 y | ↑ |
| Carotenuto et al 2020 Journal of Neurology | 10.8 y | ↑↓ |
| Van Geest et al 2018 NeuroImage: Clinical | 11.05 y | something else |
| Karavasilis et al 2019 Brain Imaging and Behaviour | 9.75 y for memory preserved, 11.14 y for memomry impaired | ↑↓ |
| Hulst et al 2015 MSJ | 11.34 y | ↑ |
| Sbardella et al 2017 MSJ | 11.45 y | ↓ |
| Bonavita et al 2011 MSJ | CI: 142.3 m, CP: 130.9 m | ↓ |
| d'Ambrosio et al 2017 HBM | 13.3 y | ↑ |
| Rocca et al 2018 MSJ | 12.1 y for whole group (11.5 y for CP, 13.4 y for CI) | ↑↓ |
| Rocca et al 2010 Neurology | SPMS: 15.5 y, PPMS: 12.7 y | ↓ |
| Meijer et al 2018 NeuroImage: Clinical | (symptom duration reported) IPS- impaired; 15.82 y, IPS-preserved; 9.8 y | ↑ |
| Eijlers et al 2018 Radiology | (symptom duration reported) CP with no atrophy 13.47 y, CI with no atrophy 13.73 y, CP with atrophy 13.61 y, CI with atrophy 16.02 y | ↑ |
| Rocca et al 2016 Brain Struct Func | MS: 13.7 y, CI: 16.1 y, CP: 12.4 y, RRMS: 9.1 y, BMS: 20.2 y, SPMS: 17.1 y | ↓ |
| Cocozza et al 2018 Journal of Neurology | 16.6 y | ↑ |
| Eijlers et al 2019 Radiology | (symptom duration reported ) CP: 13.6 y, CI 17.0 y | ↓ |
| Welton et al 2020 Brain Connectivity | 17 y | something else |
| Eijlers et al 2017 Neurology | (symptom duration reported) CP: 13.58 y, MCI: 14.15 y,, CI: 17.05 y | ↑↓ |
| Meijer et al 2017 Neurology | (symptom duration reported) CP: 10 y, MCI: 13 y, CI: 18 y | ↑ |
| Meijer et al 2018 JNNP | (symptom duration reported) early RRMS; 6.6 yyears, late RRMS; 19.0 y, SPMS; 21.8 y | ↑ |
| Fuchs et al 2019 Human Brain Mapping | 19.97 y | Something else |
| Riccitelli et al 2020 Journal of Neurology | 20.0 y | - |
| Lin et al 2019 MSJ | 21.92 y +/- 10.49 | ↑ |

* See main manuscript for full citations of studies.

†Because several studies used samples of mixed phenotypes and different disease durations, the following decisions were taken when ordering studies by the disease duration: 1) studies were ordered by the overall disease duration of the sample, when given; 2) studies were ordered by the disease duration of the cognitively impaired group; 3) if there were two cognitively impaired groups, studies were ordered by the disease duration of the more impaired group, or the cognitively impaired group with atrophy, in one case; 4) when the disease duration was only reported for each MS phenotype, or sex, studies were ordered by the disease duration of the larger sample; 5) for a study which had equal numbers of males and females, the study was ordered by the sex with the longer disease duration; 6) for one study that used a subset of MS patients that were matched to HC, the study was ordered by the disease duration of the matched subset.

‡Key: High FC is denoted by ↑; low FC by ↓; studies demonstrating both high and low FC associated with worse cognition are denoted by ↑↓; studies which did not find support for a relationship between FC alterations and worse cognition are denoted by -; “something else” refers to studies which used a methodology that does not fit into the groupings presented in this table

|  | High FC | Low FC | Negative result |
| --- | --- | --- | --- |
| Mixed phenotype sample | 13 | 16 | 2 |
| RRMS only | 11 | 10 | 3 |
| PPMS only | 1 | 1 |  |
| BMS only |  |  | 1 |
| CIS only |  |  | 1 |
| MS NOS | 1 |  |  |

**Figure 1. Tally of studies showing directional FC results, sorted by disease phenotype**

Figure 1 shows the number of studies showing high and low FC, respectively, associated with worse cognition, sorted by the MS phenotype in the sample of each study. Where studies have demonstrated both high and low FC associated with worse cognitive test performance, they have been counted twice. The total tally is therefore higher than the number of studies in the review. Studies utilising a methodology which does not show directionality of results have not been included in the figure.
